# COVID-19 in French Nursing Homes during the Second Pandemic Wave: A Mixed-Methods Cross-Sectional Study

**DOI:** 10.1101/2021.12.12.21267681

**Authors:** Morgane Dujmovic, Thomas Roederer, Séverine Frison, Carla Melki, Thomas Lauvin, Emmanuel Grellety

## Abstract

**Introduction:** French nursing homes were deeply affected by the first wave of the COVID-19 pandemic, with 38% of all residents infected and 5% dying. Yet, little was done to prepare these facilities for the second pandemic wave, and subsequent outbreak response strategies largely duplicated what had been done in the spring of 2020, regardless of the unique needs of the care home environment.

**Methods:** A cross-sectional, mixed-methods study using retrospective, quantitative data from residents of 14 nursing homes between November 2020 and mid-January 2021. Four facilities were purposively selected as qualitative study sites for additional in-person, in-depth interviews in January and February 2021.

**Results:** The average attack rate in the 14 participating nursing facilities was 39% among staff and 61% among residents. One-fifth (20) of infected residents ultimately died from COVID-19 and its complications. Failure-to-Thrive-Syndrome (FTTS) was diagnosed in 23% of COVID-positive residents. Those at highest risk of death were men (HR=1.78; IC95: 1.18 – 2.70; p=0.006) with FTTS (HR=4.04; IC95: 1.93 – 8.48; p<0.001) in facilities with delayed implementation of universal FFP2 masking policies (HR=1.05; IC95: 1.02 – 1.07; p<0.001). The lowest mortality was found in residents of facilities with a partial (HR=0.30; IC95: 0.18 – 0.51; p<0.001) or full-time physician on staff (HR=0.20; IC95: 0.08 – 0.53; p=0.001). Significant themes emerging from qualitative analysis centered on (i) the structural, chronic neglect of nursing homes, (ii) the negative effects of the top-down, bureaucratic nature of COVID-19 crisis response, and (iii) the counterproductive effects of lockdowns on both residents and staff.

**Conclusion:** Despite high resident mortality during the first pandemic wave, French nursing homes were ill-prepared for the second, with risk factors (especially staffing, lack of medical support, isolation/quarantine policy etc) that affected case fatality and residents’ and caregivers’ overall well-being and mental health.

**SUMMARY BOX:** *What is already known?:*

- Though much was learned about COVID-19 in nursing homes during the first pandemic wave (Spring 2020), descriptions of the second wave in these facilities is nearly absent from the scientific literature.
- Prior COVID-19 research in nursing homes has rarely been qualitative and has almost never interviewed care home residents themselves.
- First-wave research indicated that much stronger outbreak and infection prevention was urgently needed to bolster nursing facilities’ preparedness. Higher staff-to-resident ratios, less staff turnover, more masks, better organization, more medical support, and more epidemiological tools were found to reduce COVID-19’s impact.

*What are the new findings?:*

- Our results document a lack of preparedness for the second wave, with attack rates among staff (39% overall) and residents (61% overall) similar to levels seen during the first wave peak.
- Despite authorities’ claims to have reinforced these structures’ readiness, and despite much research into the needs in these environments, preventive measures (like strict lockdowns) remained largely unchanged and had a direct impact on residents, with 23% of COVID-positives also diagnosed with Failure-to-Thrive Syndrome.
- Qualitative results detailed how ill-suited and inflexible some preventive measures were for residents and staff alike. Participants described precarious and understaffed living and working conditions as substantial and long-standing difficulties that became critical risks during the COVID-19 outbreak, and compromised the response.

*What do the new findings imply?:*

- These results suggest that knowledge gained during the first pandemic wave was not consistently applied to care home policy or practice in France, and that these nursing homes were not always safe environments that considered residents’ mental health and well-being alongside infection prevention.
- Despite the high mortality of the first pandemic wave, French nursing homes were ill-prepared for the second. As a 5^th^ wave descends on France (albeit with much higher COVID-19 vaccination rates), applying the lessons from previous periods (especially with regard to staffing, isolation of the elderly, medical supplies, standard of care procedures) must be prioritized.

## INTRODUCTION

In France, state-funded nursing and care homes are the most common living arrangement for both independent seniors and those who need daily care and support. These institutions were deeply affected by the first wave of the COVID-19 pandemic, with an estimated 38% of all residents (247,000 cases) infected with SARS-CoV-2 and 5% (30,395) succumbing to the disease from March-July 2020. The workforce that staffs these facilities was also seriously affected, with an estimated 22% of all workers (90,000 cases) testing COVID-19 positive from late February to late May 2020 [1,2].

In October of 2020, when rising caseloads suggested a second pandemic wave, nursing homes again braced for the worst, since no vaccine was yet approved in France (this occurred in December 2020) and some variants had begun circulating. In November of that year, the non-governmental organization (NGO) Médecins Sans Frontières (MSF) began partnering with select nursing homes in Provence and Occitania provinces, in southern France, to bolster their COVID-19 prevention and care procedures in the midst of rapidly growing medical needs, strained facilities, and understaffing (often aggravated by absenteeism spurred by workplace-acquired infections). As nursing homes transformed into places providing hospital-level care, staff were required to perform more advanced technical procedures and increased disease surveillance at a moment when human resources were depleted due to illness and overwork. Concurrently, health authorities recommended strong lockdown measures for elderly care home residents, including bans on going outside, prohibiting family visits, and confining residents to their rooms.

Despite the devastating mortality rates seen in care homes around the world throughout the pandemic, scientific literature has not yet described the second wave of COVID-19 in this environment. Published research is mostly focused on the first pandemic wave period, almost exclusively quantitative studies or systematic reviews on specific topics. Several articles report best practices for infection prevention and control (IPC) (i.e. frequent testing for staff, residents, and visitors, staff cohorting, and strict isolation policies), or recommended better evaluation of the consequences of lockdown restrictions [3-13,14, 15]. Other lessons from the initial crisis period were that more staff [6,8], support [8,9], protective equipment, and overall preparation [8-10] could prevent or reduce outbreaks. Lately, articles focused only on impact of vaccination on transmission among staff and residents [16,17]. The little qualitative research conducted during the first wave was almost never able to conduct in-person interviews [12, 18-24], but found that lockdowns had a significant and deleterious impact on residents, staff well-being, and staff turnover [20,21].

Our research attempts to understand the risk factors that influenced the second pandemic wave, the impact of that wave, and how staff and residents experienced this period of the pandemic in a nursing home setting.

## METHODS

In this mixed-methods, cross-sectional study, we analyze retrospective COVID-19 data from 14 nursing homes being reinforced by support from MSF in order to assess the impact of the second pandemic wave as well as the effects of prevention measures on resident mortality and comorbidity. These results are given depth and detail through qualitative investigation into staff and resident experiences.

### Definitions

*Autonomy Evaluation Score* (AES) measures a care home resident’s level of autonomy. An AES of 1 reflects the lowest level of autonomy (i.e. confinement to a bed or armchair, serious mental function impairment, continuous caregiving required), while an AES of 6 refers to people who have fully retained their autonomy in their daily lives. *The Average Weighted Autonomy Score (AWAS)* is the overall AES score for a facility. This score is a proxy for the financial and human resources that a nursing home needs and has access to: the higher the AWAS, the more resources needed (staff-to-residents ratio, equipment, etc.) and the more dependent the residents. *Failure to Thrive Syndrome (FTTS)* is specific to elderly individuals and characterized by a rapid deterioration after a physical or psychological event (Appendix 1).

### Study Design and Population

This cross-sectional, mixed-methods study used retrospective, quantitative data from residents living in 14 nursing homes between November 2020 to mid-January 2021. Four nursing facilities were purposively selected as qualitative study sites for additional in-person, in-depth interviews conducted between January and February 2021. Qualitative study sites were selected based on whether they had passed their epidemic peak, had high attack and fatality rates, were public or private facilities, and their geographic location.

### Data Collection

Facilities’ administrative data (number of beds and staff, staff-to-resident ratios, etc), COVID-related data (confirmed cases among residents and staff, episode duration, deaths, etc) and individual, anonymized patient data (demographics, comorbidities, etc) were used for quantitative analysis.

Qualitative data was gathered using semi-structured, in-depth interviews (IDIs) during one-week ethnographic immersions in each of the four qualitative study sites. The lead investigator targeted four groups of actors, including facility administrators (directors, coordinating physicians, and nurses), clinical and facilities staff (nurses, caregivers, educators, physical therapists, maintenance crews), the residents themselves, and the residents’ visiting family members. Participants were purposively selected to obtain a maximally heterogenous sample of interview participants and reflect the spectrum of opinions and experiences of everyday life nursing homes. Across the 4 qualitative study sites, a total of 47 IDIs were conducted with facility directors (4), staff members (36), and residents (7). Among the 36 staff members, 29 were caregivers and 7 provided other support functions (human resources, maintenance, cleaning, cooking). All interviewed residents were women, as were the majority of study participants overall (82.9%). Interview length varied from 12-171 minutes (54-minute average) (Appendix 4).

Telephone and face-to-face interviews were also conducted with 10 residents’ family members, though family interviews are not included here in order to focus on experiences from within the nursing homes during lockdown. Nine residents refused to participate (due to fatigue, discomfort with interviewing, or COVID-19 related reasons). Caregiver participation was constrained by understaffing, overwork, fatigue, or disease, which left them with very little time or energy for interviewing.

Vulnerable residents were pre-selected under the advisement of the coordinating nurse on the permanent caregiver teams. Participants had to be able to give informed consent, capably interact, and have no major cognitive disorders. Level of autonomy (AES) did not constitute an a priori criteria for participant selection. Whenever a legal guardian or curator was designated, the latter was contacted prior to the interview to verify that consent could be obtained from the interviewee.

Question guides focused on three primary topics: the outbreak chronology, adaptation to the crisis, and the individual experience of the second pandemic wave (Appendix 3). Individual guides were adapted for those living in the nursing home (residents) or working there (facility administrators and staff). All interviews were voice recorded and direct observations were written in the investigator’s field book. All written data were anonymized upon collection. Participants’ personal data was assigned a study number that was set on a correspondence table kept separately from other data. Written informed consent was obtained prior to each interview.

Preventive measures were implemented with all participants to decrease COVID-19 disease transmission risk: systematic FFP2 face mask use, social distancing, hand and space disinfection, and weekly Rt-PCR tests for the two field investigators.

### Statistical Analysis

Patient data were explored using univariate analysis to highlight possible mortality risks. Univariate unadjusted Cox Hazard Ratios, Kaplan-Meier estimations, and Log-Rank tests were used for multivariate analysis. A stepwise procedure was followed, retaining factors with a log-rank test value <0.3. COVID-19 mortality was estimated using a multilevel mixed-effects Cox model using selected factors identified in the univariate analysis. Random effects on individual variables were considered and nested at the facility level [25]. Interactions between potentially correlated factors (comorbidities, failure-to-thrive syndrome, autonomy level, time-related variables) were accounted for while robust standard errors were computed (Appendix 7). 95% confidence intervals are presented and a significance threshold of 5% was chosen for p-values. Statistical analyses were conducted with Stata 15^®^ and R Studio 1.4^®^.

### Qualitative Analysis

Data analysis was performed from January to March 2021, similar to the fieldwork period (January-February) and reporting phase (March-April). Qualitative analysis combined grounded theory and hypothetico-deductive analysis. Preliminary observation in five nursing homes and MSF-team reports were used to create an initial checklist for systematic direct observation. In January and February 2021, 36 semi-structured IDIs were conducted in three nursing homes, in combination with “external participatory observation” [26]. Questions were adjusted in an iterative manner after preliminary analysis was conducted on these initial interviews. Data saturation was sought throughout the interview process and discussed within the research team on a weekly basis. In February 2021, 11 semi-structured IDIs were conducted in a fourth nursing home in order to assess data saturation.

Interview data were processed gradually through professional transcription and verified with the interviewees when necessary. De-identification occurred during transcription (names, places, dates, distinctive personal data etc). Interview data were written, analyzed and coded in Excel spreadsheets. A first codebook with 39 data codes emerged from interview transcripts. Five themes were initially analyzed and refined into a final set of 33 across four key categories. Three of these were cross-cutting and had up to three sub-themes (Table 3). Results are reported in accordance with the Standards for Reporting Qualitative Research (SRQR) guidelines [27] and the COnsolidated criteria for REporting Qualitative research (COREQ) checklist.

### Patient and Public involvement

Administrators and coordinating physicians from 14 nursing homes were actively involved in collecting and anonymizing study data from their residents/patients. During the exploratory phase of research (December 2020 to January 2021), any feedback from qualitative study site administrators was included in the study protocol. During data collection (January to February 2021), the research methodology was discussed with MSF nurses and facilities staff and adapted to each nursing home’s context and caregiver guidance. At the beginning of each IDI, caregivers and residents were encouraged to further participate in the research by contacting the lead investigator with any suggestions. In the reporting phase (from the 1st March to June 2021), internal reporting was sent to interviewees who wanted to be contacted for this purpose. This report was sent to prominent political COVID-19 crisis management actors (such as the French Ministry of Health). A summary letter will be brought to resident study participants and facility staff to inform them of the results and gather their comments on possible follow-up.

### Ethics

This study received approval from the MSF Ethical Review Board (ERB) and the Commission Nationale de l’Informatique et des Libertés (CNIL) in France. Patient data and qualitative observations were fully anonymized. All study procedures were in line with the Declaration of Helsinki.

## RESULTS

22 nursing homes were originally included in the study, though data was available for only 14 of them (the others did not send data in time for analysis or the data were not electronically recorded). The 14 participating nursing facilities were largely state supported entities (79%) with an average of 68 residents (median=65; IQR: 58-73). Results varied considerably from one nursing home to another. COVID-19 outbreak duration averaged 39 days (median=40; IQR: 30-50 days) while Infected residents’ individual COVID-19 episodes averaged 24 days (median=30; IQR: 14-51 days). The average attack rate was 39% (median=39%; IQR: 29%-54%) among staff and 61% (median=60%; 50%-73%) among residents. One-fifth (median=20%; IQR: 17%-23%) of the residents who were infected ultimately succumbed to COVID-19 and its complications. The mean Average Weighted Autonomy Score (AWAS) was 770 (median=763 ; IQR: 722-804) and the average staff-to-resident ratio was 0.82 (median=0.86 ; IQR: 0.72-0.90). The average time to universal masking policies being implemented was 9.6 days (median=6.5 ; IQR: 2-15 days) and average time until a facility was bolstered with MSF support (staff or resources) was 17.5 days (median=15; IQR: 13-28 days). (Appendix 4).

### Patient Risk factors

Retrospective COVID-19 data were obtained for 14 nursing homes, finding 585 COVID-19 cases among 930 residents (61% attack rate) (Table 1). Cases were mostly women (78%) who were >85 years old (68%). Individual Autonomy Scores (IAS) were low (<2) in a majority of cases (60%), indicating a very low-level of autonomy overall. One-fifth (21%) of cases were transferred to a hospital, while half (46%) were put on oxygen therapy. One-tenth (12%) of COVID-cases received palliative care, and nearly one-quarter (22%) died. Failure-to-Thrive Syndrome was diagnosed in nearly one-quarter (23%) of COVID-positive residents. At least one other comorbidity was found in over half (61%) of infected residents. AWAS, nursing home size, and staff-to-resident ratios were all strongly correlated, as were time-related variables (time until external MSF support was received, time until universal masking policies were applied, and duration of COVID episode) (Table 1).

**Table 1.**
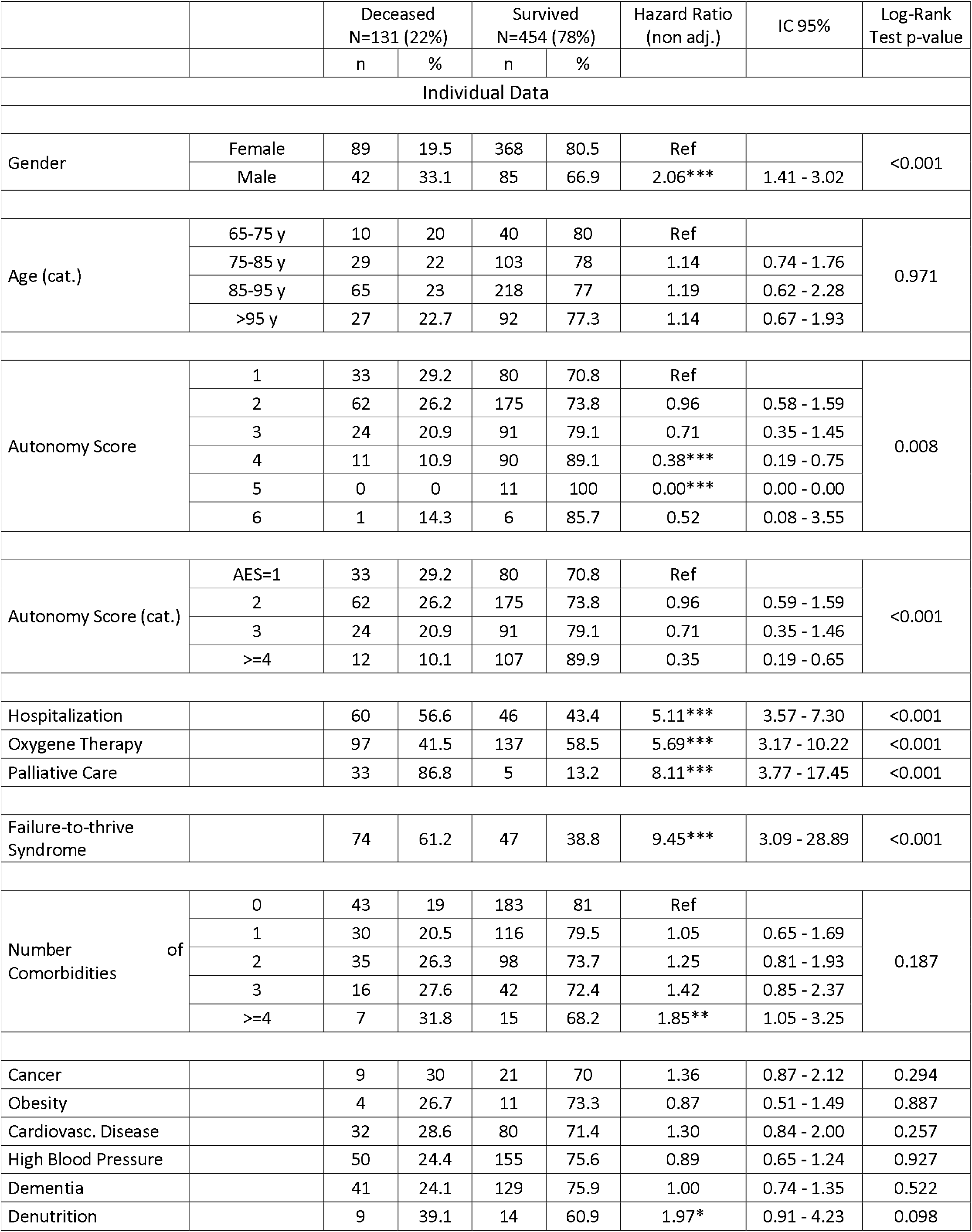
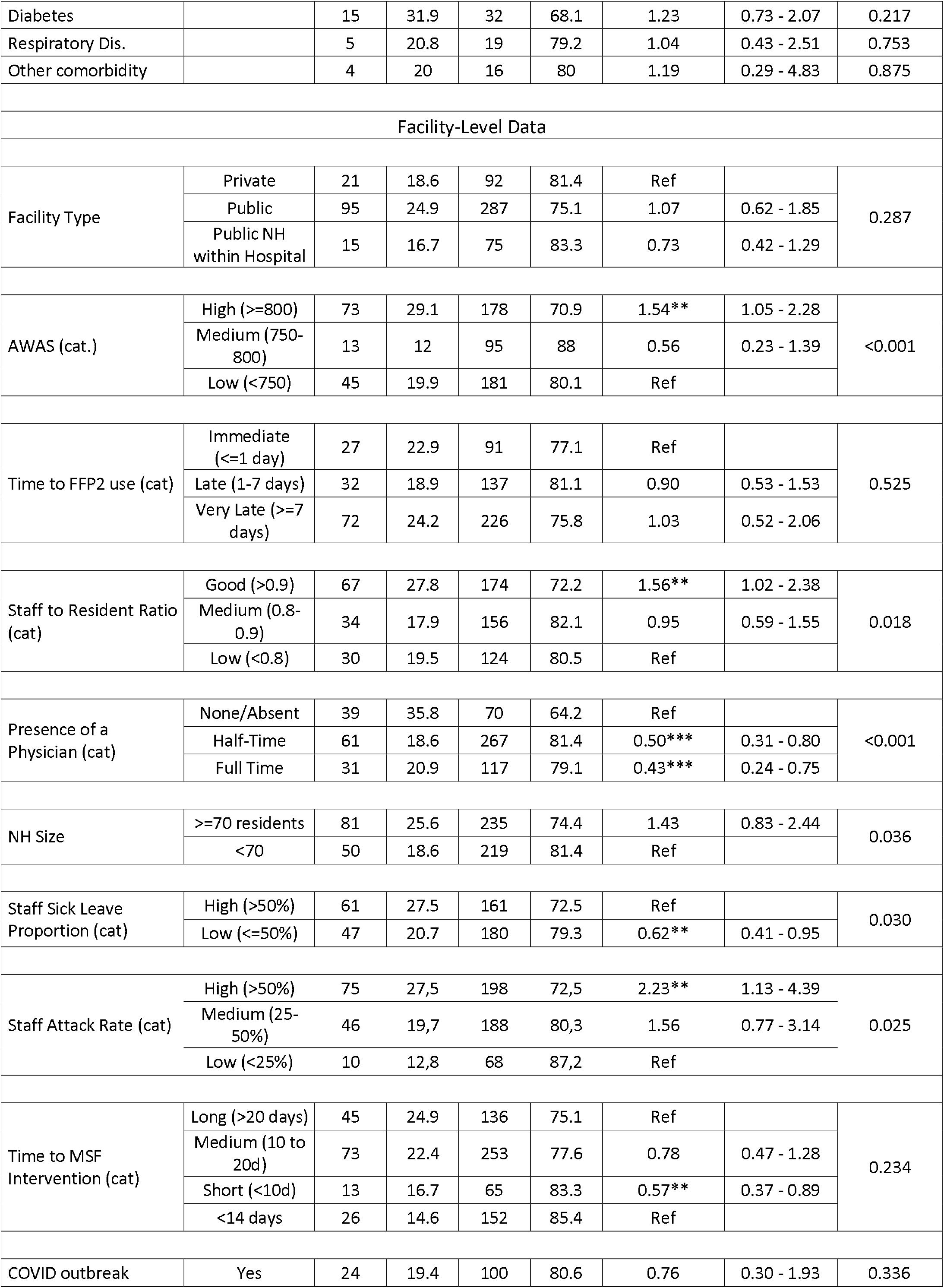
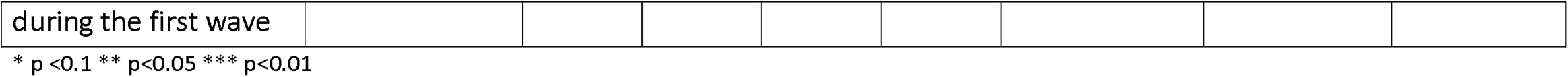
Univariate Analysis of Nursing Home Resident and Facility Data, Provence and Occitania Provinces, France, 2021

Univariate analysis using Cox modelling (Table 1) and Kaplan-Meier estimations (Figure 1) suggested that individual characteristics like gender (log rank p<0.001) and IAS (p=0.008) were associated with COVID-19 mortality, while age and specific comorbidities were not. Survival curves also suggested that facility characteristics like low AWAS (p<0.001), the absence of a permanent physician on site (<0.001), larger nursing home size (>70 residents) (p=0.036), and a high staff attack rate (p=0.025) were also associated with resident mortality. Predictably, hospitalization (p<0.001), palliative care (p<0.001), and oxygen therapy (p<0.001) were all strongly correlated with the risk of death, as was the presence of FTTS (p<0.001) and the presence of more than 4 co-morbidities (risk increased with the number of co-morbidities present, p=0.045). Additional Kaplan-Meier Curves for non-significant factors can be found in the supplementary information (Appendix 5).

**Figure 1.**
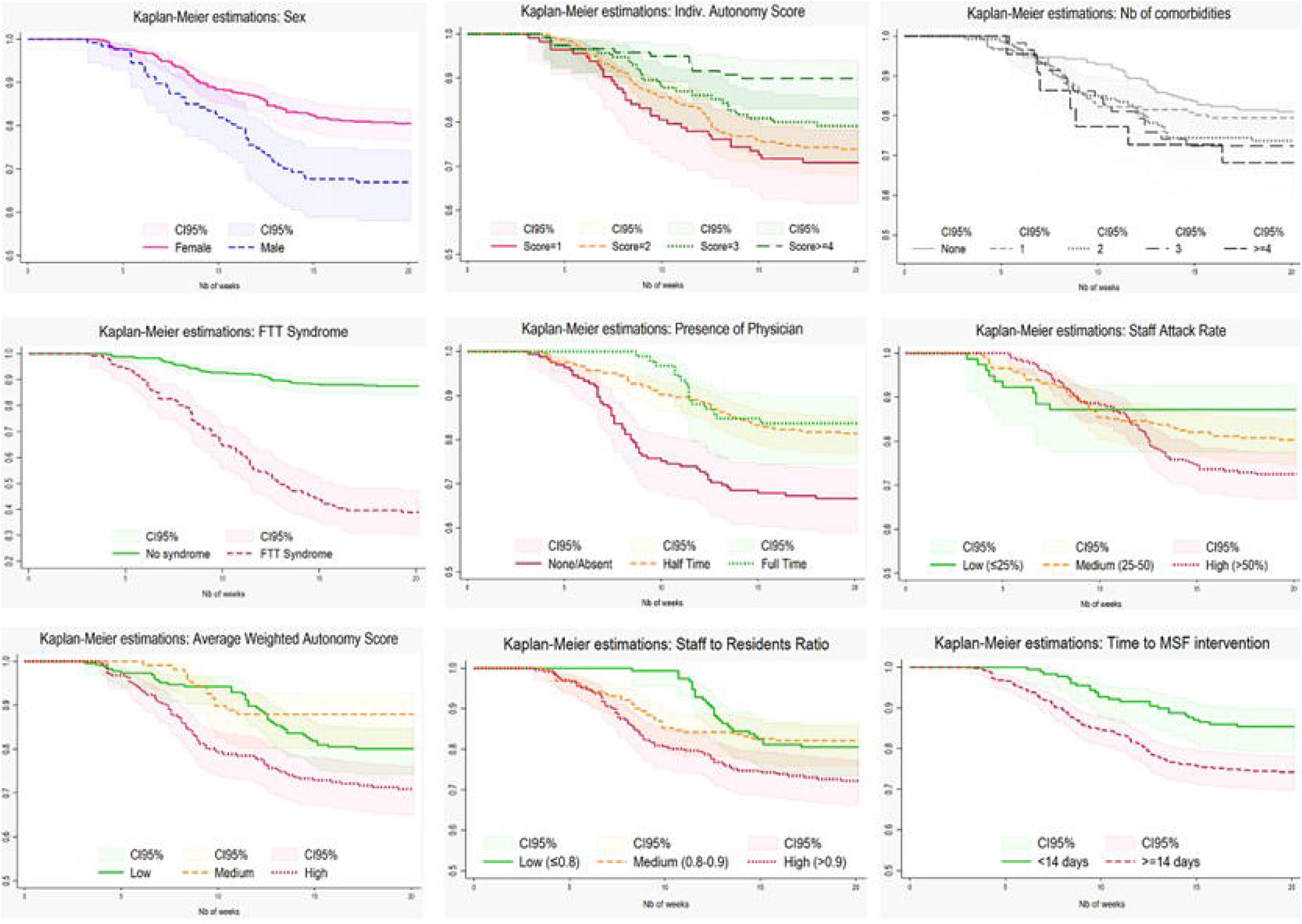
Likelihood of survival by resident and nursing facility characteristic, Univariate (Kaplan-Meier) Analysis, Provence and Occitania Provinces, France, 2021. On the X axis: Number of weeks from Oct 15^th^, 2020 ; On the Y axis: probability of resident’survival.

Multilevel Cox Hazard modelling highlighted mortality associated factors adjusted for potential confounders (Figure 2). Those at highest risk of death were men (HR=1.78; IC95: 1.18 - 2.70; p=0.006) with an FTTS diagnosis (HR=4.04; IC95: 1.93 - 8.48; p<0.001) in facilities with delayed implementation of universal masking policies (HR=1.05; IC95: 1.02 - 1.07; p<0.001). The lowest mortality risk was found in residents of facilities with a partial (HR=0.30; IC95: 0.18 - 0.51; p<0.001) or full-time physician on staff (HR=0.20; IC95: 0.08 - 0.53; p=0.001), with individual AES scores >3 (HR=0.38; IC95: 0.16 - 0.89; p=0.026). Noticeably, higher AWAS (a proxy for staff-to-resident ratios and a nursing home’s overall means) was associated with a lower risk of death (HR=0.99; IC95: 0.99 - 1.00; p=0.020) (Table 2). Sensitivity analysis can be found in the supplementary information (Appendix 6).

**Figure 2.**
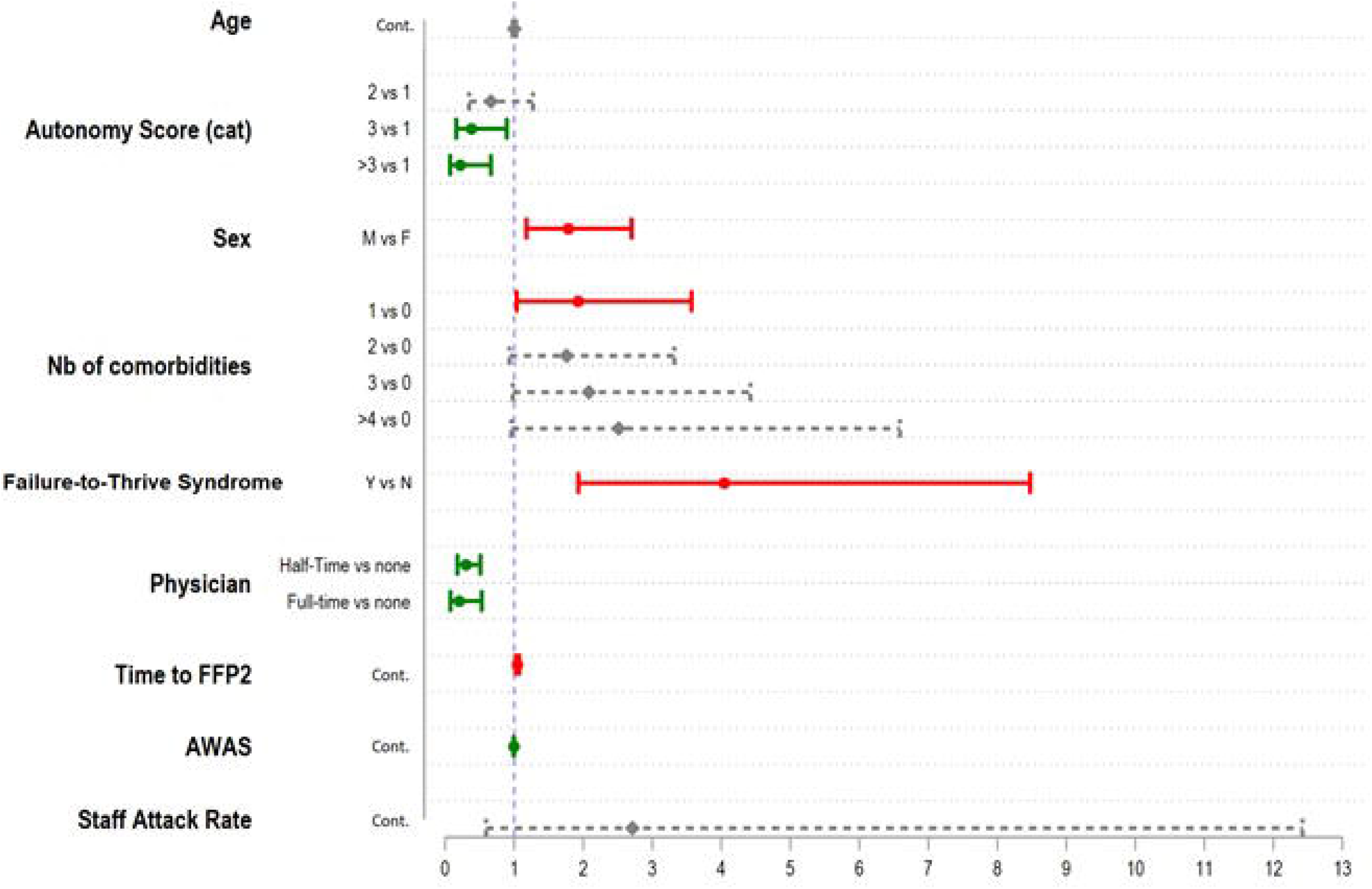
Final Cox Model: Forest Plot of mortality associated factors in French nursing facilities, Provence and Occitania provinces, 2021. (HR>1), full-lines in green for protective factors (HR<1) and dashed-lines in grey for CI95% of non-significant factors.

**Table 2.** Multivariate Cox Hazard adjusted analysis of mortality associated factors in French nursing facilities, Provence and Occitania provinces, 2021 (Information Criteria: AIC*=1171; BIC=1226)

| VARIABLES |  | Adjusted Hazard Ratio | CI95 | p-value |
| --- | --- | --- | --- | --- |
| Age | Continuous | 1.00 | 0.98 - 1.03 | 0.876 |
| Autonomy Score | 2 vs 0 | 0.66 | 0.35 - 1.27 | 0.216 |
|  | 3 vs 0 | 0.38 | 0.16 - 0.89 | 0.026 |
|  | ≥4 vs 0 | 0.22 | 0.07 - 0.66 | 0.007 |
| Gender | M vs F | 1.78 | 1.18 - 2.70 | 0.006 |
| Comorbidities | 1 vs 0 | 1.92 | 1.04 - 3.57 | 0.038 |
|  | 2 vs 0 | 1.76 | 0.93 - 3.32 | 0.081 |
|  | 3 vs 0 | 2.08 | 0.98 - 4.42 | 0.056 |
|  | >=4 vs 0 | 2.51 | 0.96 - 6.59 | 0.061 |
| Failure-to-thrive Syndrome | Y v N | 4.04 | 1.93 - 8.48 | 0.000 |
| Presence of a physician | Half Time vs None/Absent | 0.30 | 0.18 - 0.51 | 0.000 |
|  | Full Time vs None/Absent | 0.20 | 0.08 - 0.53 | 0.001 |
| Time to FFP2 use (in days) | continuous | 1.05 | 1.02 - 1.07 | 0.000 |
| AWAS | continuous | 0.99 | 0.99 - 1.00 | 0.020 |
| Staff Attack Rate (%) | continuous | 2.71 | 0.59 - 12.42 | 0.198 |
| Interaction Terms |  |  |  |  |
|  | AES=2#FTTS=1 | 2.26 | 0.90 - 5.67 | 0.083 |
|  | AES =3#FTTS=1 | 3.10# | 1.00 - 9.58 | 0.050 |
|  | AES =4#FTTS=1 | 4.79# | 1.16 - 19.87 | 0.031 |

**Table 3.**
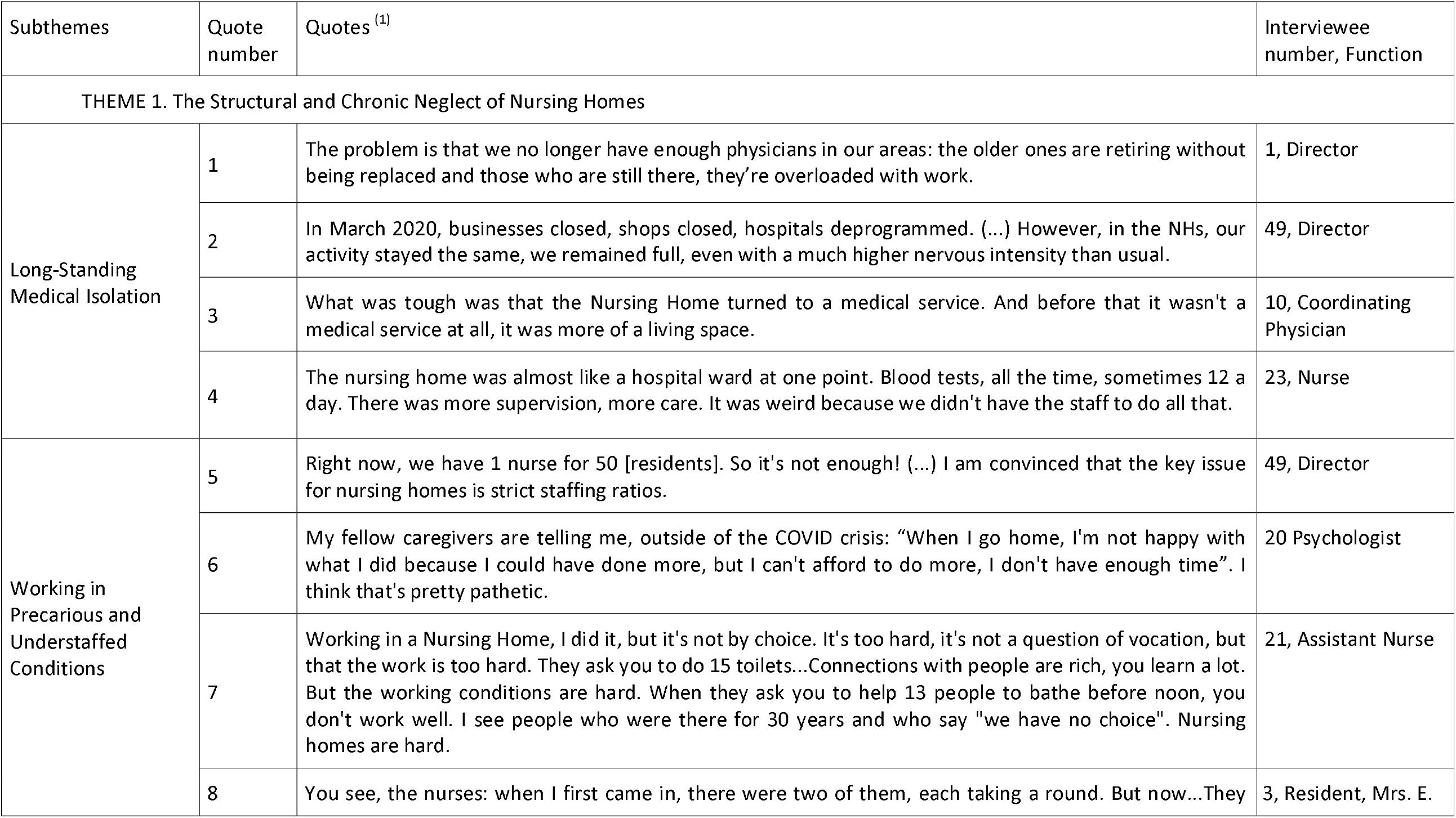
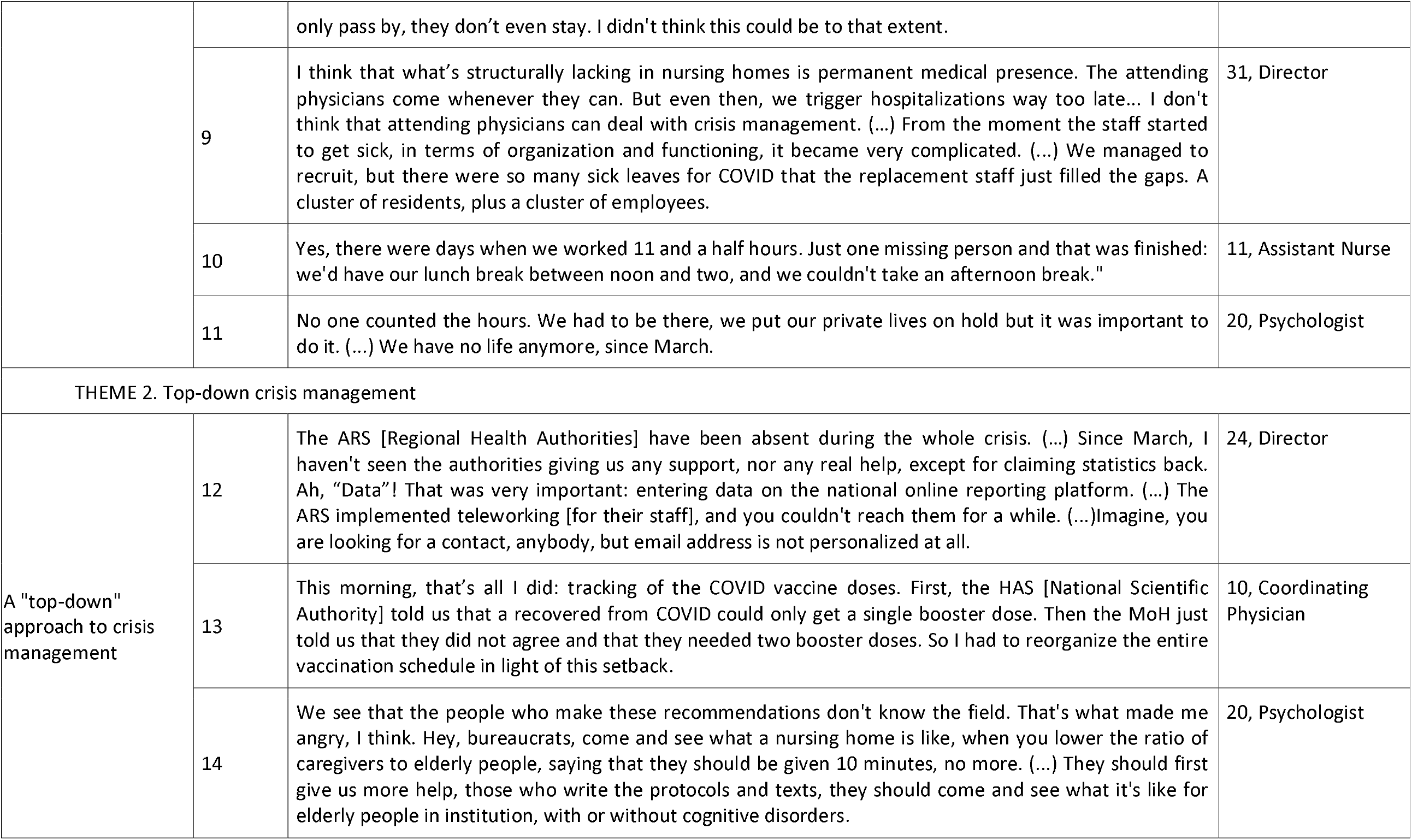
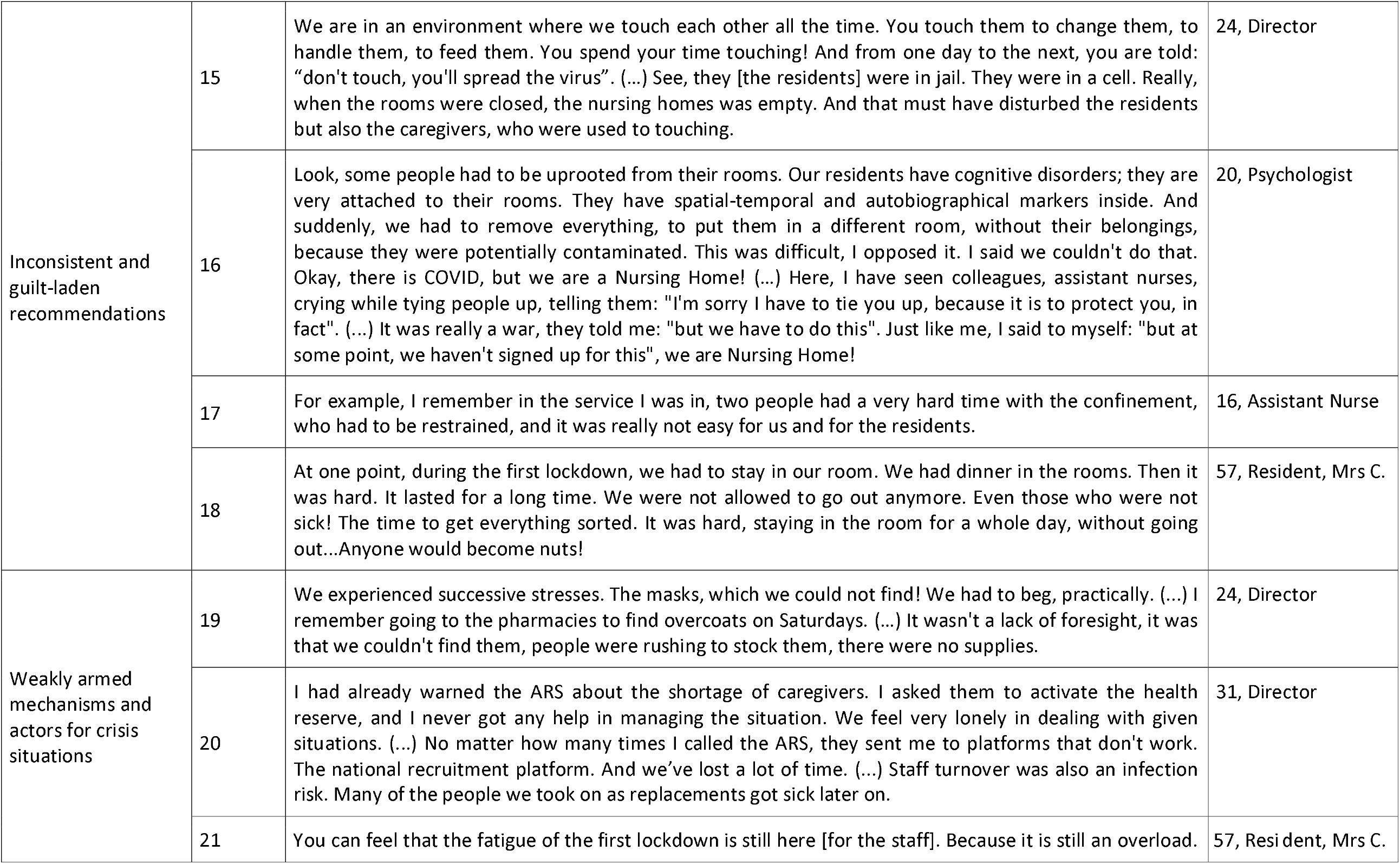
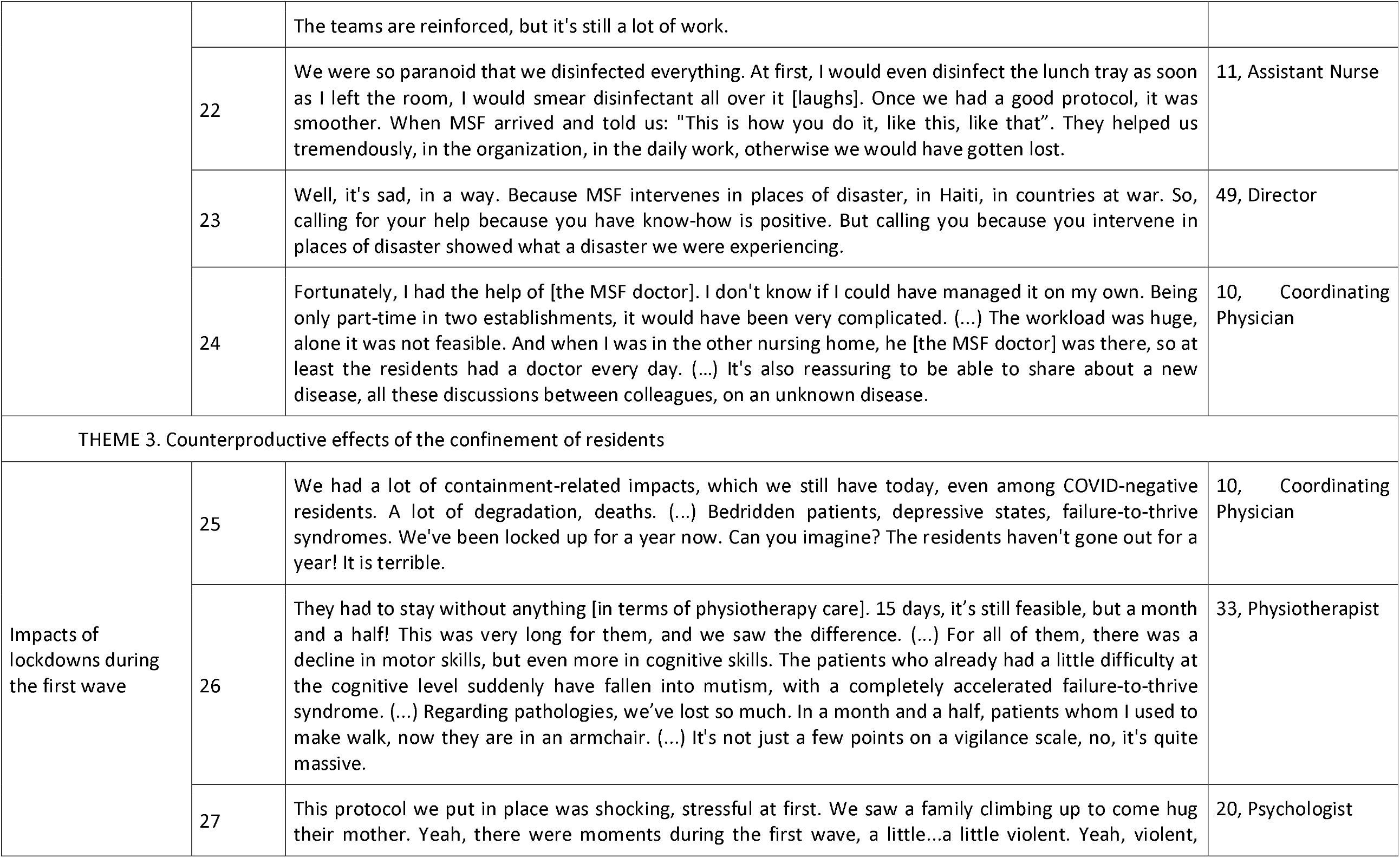
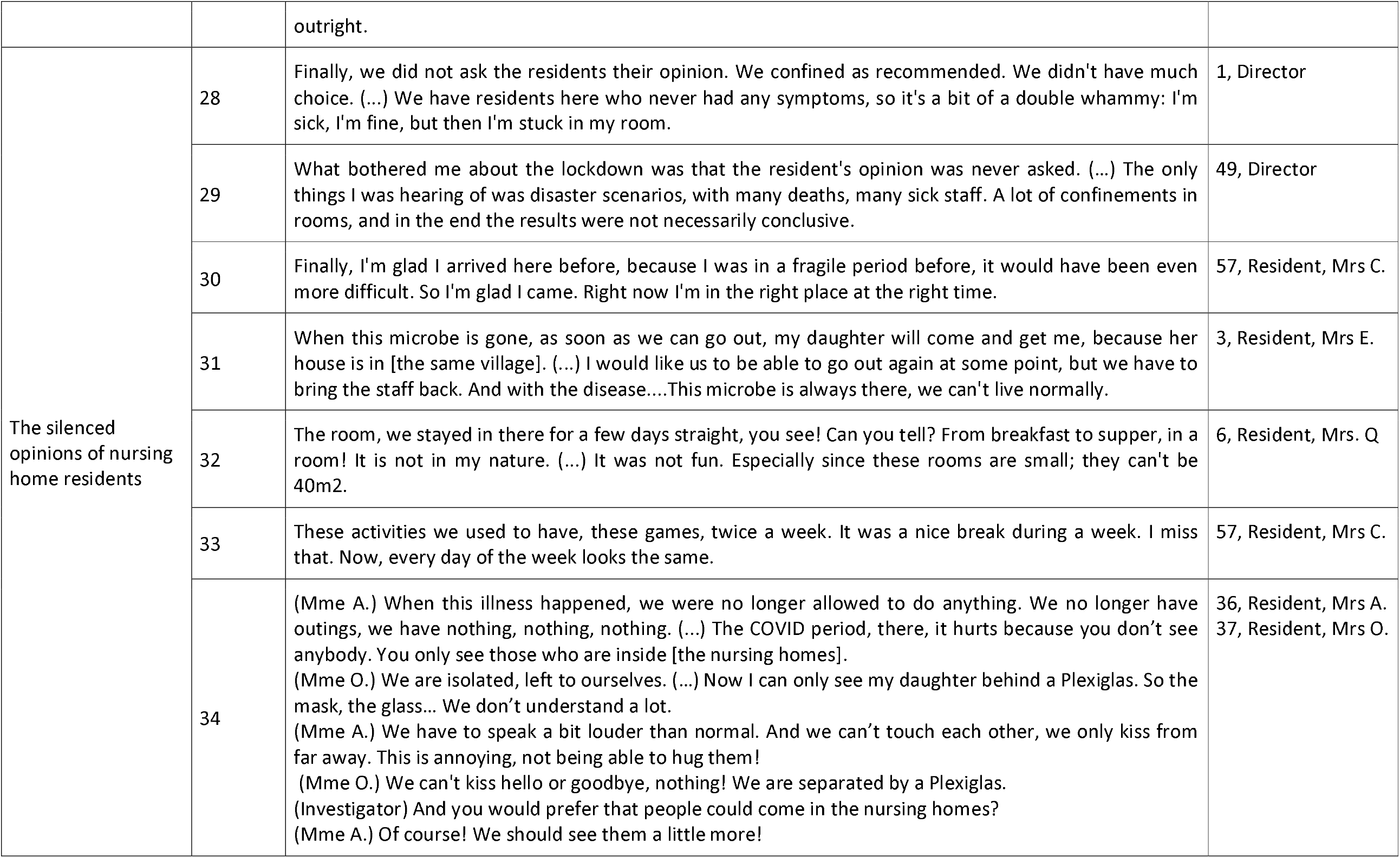
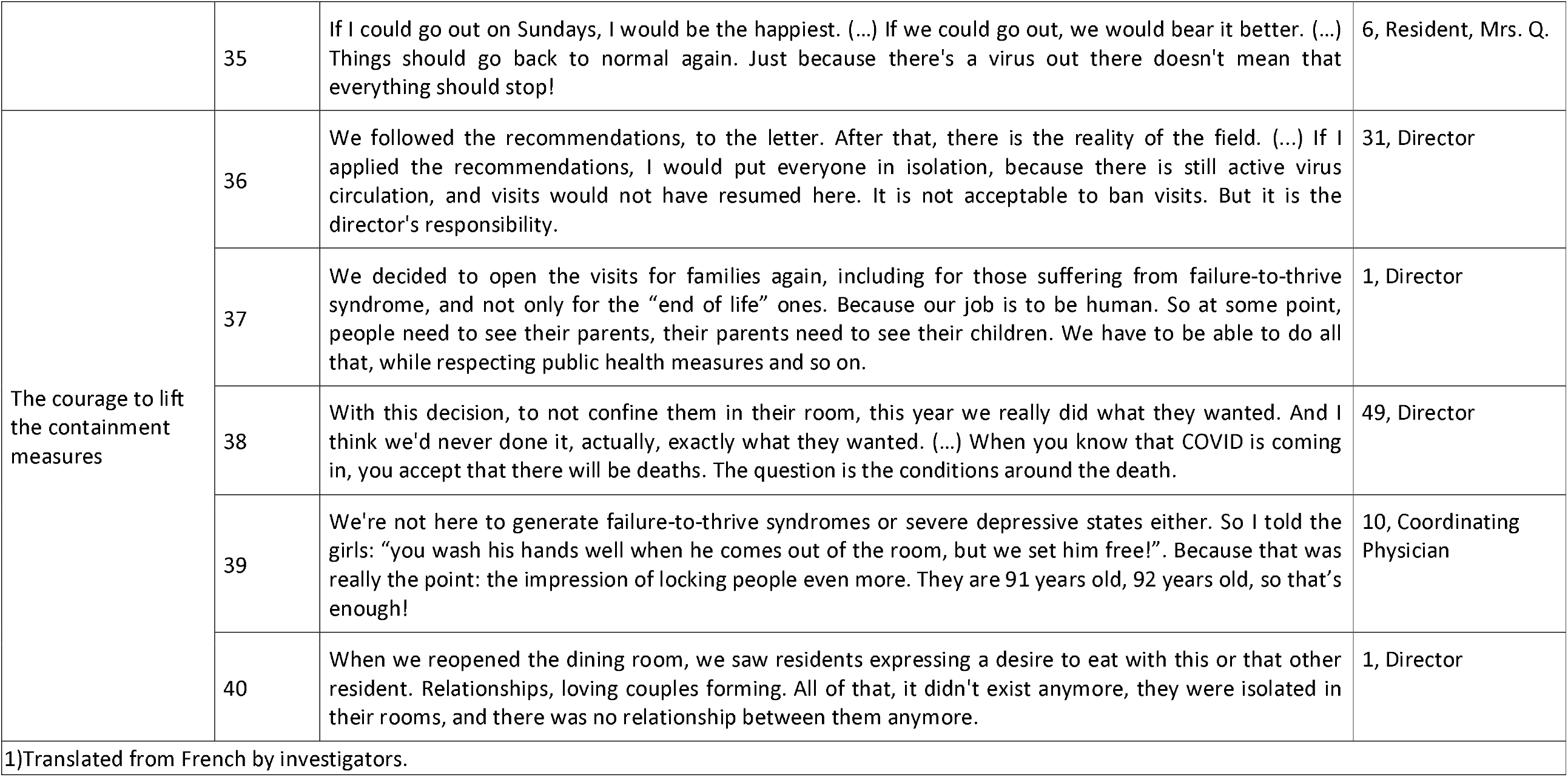
Representative quotes for the 3 themes

### Qualitative Results

The qualitative approach richly described interviewees’ lived experiences during the COVID-19 crisis, revealing difficult-to-quantify social influences on the outbreak’s evolution and impact. Three significant themes emerged from our discussions (Table 3).

#### Structural, Chronic Neglect of Nursing Homes

Staff members described a long-standing lack of physicians in nursing homes, exacerbated by lockdowns and growing medical needs during a period of rising COVID-19 infections. One nurse explained, “the nursing home was almost like a hospital ward at one point…There was more supervision [needed], more care…We didn’t have the staff to do all that.” All groups of interviewees emphasized that working in precarious and understaffed conditions was a substantial difficulty that became a critical risk during the COVID-19 outbreak and compromised the response. Assistant nurses described extremely challenging working conditions: “When they ask you to help 13 people to bath before noon, you don’t work well.” This situation was worse during the second pandemic wave when, as one psychologist explained, “no one counted the hours. We had to be there, we put our private lives on hold, but it was important to do it.” All directors described a structural lack of a “permanent medical presence” and the need of a “strict staffing ratio.”

#### Top-down Crisis Management

Personnel highlighted the “top-down” approach of French health authorities, including a lack of communication and time-consuming processes for staff and administrators alike, “The ARS [Regional Health Authorities] have been absent during the whole crisis. (…) Since March, I haven’t seen the authorities giving us any support, nor any real help, except for claiming statistics back.” These officials worked far from the frontline environment of a nursing home and were removed from the suffering of residents and staff. As a result, it was felt that they encouraged ill-informed, unrealistic, and inconsistent crisis-response measures: limiting contact with residents, confining them to their (small) rooms, abruptly relocating them in new rooms (very disturbing for them) or even physically restraining residents in distress. A psychologist described how “some people had to be uprooted from their rooms” where they had “spatial-temporal and autobiographical markers”, while others “had to be restrained” by assistant nurses. All of these were deeply disheartening to staff and residents, creating feelings of shame and guilt among caregivers and the potential for cognitive disorders among residents. A resident explained that “it was hard, staying in the room for a whole day, without going out,” and that “anyone would become nuts!” Weak crisis response mechanisms also manifested as poor prevention measures (a lack of universal masking requirements initially, facemask shortages during the first wave), lack of state medical relief staff, and such an extreme lack of preparedness that assistance from a non-state humanitarian actor like MSF was needed. As a director told us, calling MSF, a disaster-response organization “showed what a disaster we were experiencing.”

#### Counterproductive Effects of Lockdowns

Finally, participants described the counterproductive effects of lockdowns, including negative medical outcomes and even violence. Physiotherapists described “a decline in motor skills, but even more in cognitive skills” and “completely accelerated failure-to-thrive syndrome” which corroborates other descriptions of “bedridden patients, depressive states, failure-to-thrive” because “the residents haven’t gone out for a year.” Participants were discouraged that lessons from the first pandemic wave did not translate into better preparedness and smoother, more nuanced, and less restrictive lockdown policies during the second. Despite feeling secure in their nursing home environment during the pandemic period, interviews with residents revealed the depth of their dislike for the extreme physical and social isolation they faced while alone in their rooms, especially when facilities’ social activities, family visits, and outings were suspended or strictly supervised with social distancing measures. Extreme fatigue occurred after a year of lockdown and social restrictions, as one nursing home’s 90-year-old resident explained “if we could go out, we would bear it better.” Since facility administrators were urged to follow the ARS recommendations, only a few directors or staff were willing to soften lockdown measures, allow family visits, or take residents’ end-of-life wishes or needs for social interaction into account.

These interviews show some overlap with the risk factors that were highlighted in the quantitative data (mortality risks linked to understaffing, the absence of a permanent staff physician, low staff-to-resident ratios, lockdowns linked to FTTS). Other qualitative factors associated with better pandemic management also appeared in interviews, such as reliable communication with local health authorities, the presence of an effective national health strategy, and collaboration with other medical sectors.

## DISCUSSION

Our study is the first mixed-methods investigation of nursing homes during the COVID-19 pandemic in France, and one of the first in Europe. MSF staff’s close, in-person work with these care facilities gave investigators privileged access during a challenging period and lead to particularly rich interviews. This lies in contrast to most other qualitative investigation of the geriatric population during the COVID period, which have usually been conducted remotely or via surrogates (caregiving staff or family members), without being able to interview residents themselves. These results show clearly that the second wave looked largely similar to the first wave in French nursing homes, in both response and impact, and that these facilities were not sufficiently prepared and supported when facing subsequent threats to their vulnerable tenants.

Nursing home data is not routinely collected by French national health information services because residents are considered to “live at home.” Thus, considering how difficult it is to access even the most basic data from these facilities (such as number of cases or deaths), we managed to construct a large dataset containing detailed information about COVID-19 cases, which affected 30% of all residents in the 14 participating nursing homes. The study also allowed a thorough examination of COVID-19 as experienced by the staff and residents who most suffered from the pandemic. To the best of our knowledge, French crisis management measures during the second pandemic wave were never informed by qualitative data. In this study, patients’ risk factors could be explored in relation to influential social and structural determinants of health, such as understaffing, strict lockdown measures, isolation from other medical actors/lack of medical support or the top-down and bureaucratic crisis management by health authorities.

Our multivariate analyses confirmed mortality trends seen in other settings. Similar to other studies, we found that men died more often despite their being a minority of nursing home residents, and that residents’ autonomy was a strong factor in their survival, with those who were more reliant on staff for daily support most likely to succumb to their disease [8-10, 28-32]. Living with multiple comorbidities (especially diabetes and dementia) was also strongly predictive of COVID-mortality in our group [8, 10, 28-32]. The negative effects of understaffing (seen as sick leave or AWAS in our data) were similar to those reported in the United States [8], Spain [32], and the United Kingdom [33-34], and constitute a vicious cycle: during periods of high transmission, more staff needed sick leave. Yet, the medical and staffing needs of residents were simultaneously surging, forcing many sick (and infectious) caregivers back into the workplace. The cycle was compounded by the destructive effects that an enormous workload and an anxiety-producing work environment is known to have on caregivers’ wellbeing [12, 18, 20, 21, 36].

The efficacy of universal masking to prevent respiratory disease is well established [37,38], though we were not able to measure the impact of staff/resident masking because mask mandates were often put in place at the same time that extra resources and support from MSF arrived and bolstered the nursing facility overall. Nevertheless, our results do suggest that higher transmission and case fatality were associated with delays in mandatory mask requirements for staff, confirming the utility of these rules in uniquely vulnerable and high-risk nursing home settings. The facemask issue is not easy, however, in a nursing home context. The health benefits of masking have trade-offs with other social needs: care home residents may live with hearing or cognitive disorders, and masking may prevent voice and facial recognition or communication. The absence of others’ daily smiles or expression may have led to cognitive decline, a point that has been shown in previous research and was emphasized in our interviews with caregivers, managers, and residents alike [39, 40].

Finally, the benefit of confining residents to their rooms is strongly questioned by these results. While such measures undeniably reduce virus transmission among residents [6-10, 14-15, 33, 37-38, 41-43]; the consequences for their mental health and nutritional status have also been shown to be considerable [12, 13,20-24, 36, 44-48]. Strict lockdowns in our cohort were associated with higher FTTS incidence, triggered by individuals’ difficult living conditions over multiple months (the long duration of the crisis, an anxiety-provoking atmosphere, social isolation, other residents’ deaths, etc.). We found a strong statistical association between COVID-19 case fatality and FTTS diagnoses, a result that was triangulated by qualitative interview data and is consistent with other research from France [41], the United Kingdom [42], Finland [46], Spain [48,50], Italy [49], and the United States [47].

### Limitations

Our study is limited by the fact that study site selection was not random but was instead steered by discussions with MSF. Moreover, since MSF targeted mostly struggling nursing homes, the study included only a small number that did not have major outbreaks (or contained their outbreaks early). As a result, comparing these facilities to others in Provence and Occitania (or France) should be made with care. Participant selection was biased by the fact that only residents who were fully capable of interacting with investigators and were able to give informed consent could be interviewed, thus excluding anyone with major cognitive disorders (a relatively frequent condition in nursing homes). Quantitative data were neither exhaustive nor always electronically recorded. Associations between COVID-19 deaths and FTTS were complicated by the co-morbidities that many residents also lived with, though adjusted analysis attempted to control for potential confounding.

## CONCLUSION

These results raise questions about French health authorities’ approach to managing the second wave of the COVID-19 pandemic, as seen through the lens of those living through the crisis. If institutional management of older ager, loss of autonomy and end of life is a chronic issue for a long time in France, solutions exist to support nursing homes in times of acute crisis. Future debates about pandemic response in this setting should take into account things like the social needs of residents, understaffing as a risk factor for higher COVID-related deaths, and should refine general health policies and prevention measures in nursing homes.

Moreover, once an outbreak has occurred, tough questions must be asked: Are restrictive measures for all residents worth the personal and mental health toll? How can facilities improve residents’ end-of-life conditions in a controlled, safe way that will allow them (and their families) dignity and care? Is this reasonable to do if it involves a modicum of increased risk exposure for the facility overall? These results remind us that effective COVID-19 response should be context-adapted, patient-centered, and humane.

## Data Availability

All data produced in the present study are available upon reasonable request to the authors

## CONTRIBUTORS

CM, TR, MD, TL and EG conceived the study (literature search, study design, etc). MD, SF, TR, CM, TL and EG developed the study protocol. MD performed field data collection (qualitative interviews) and SF collected epidemiological data. TR and SF performed data management and statistical data analysis. MD performed interview transcription and qualitative analysis. MD and TR performed literature search and wrote the first version of the manuscript. TR and EG verified the underlying data and performed additional analyses. All authors interpreted the results, contributed to writing the manuscript, and approved the final version for submission.

## ACKNOWLEDGMENTS

First and foremost, authors are very grateful and thank Janet Ousley for her help on article editing. Authors also thank Marie Thomas, Tommaso Fabbri, Klaudia Porten, Michel-Olivier Lacharité, Marc Gastelly-Etchegorry, and the whole MSF team in the field. This study would not have been possible without the collaboration of the nursing home managers, staff, and residents. A very special thanks go to each and every one of them.

## FUNDING AND ALL OTHER REQUIRED STATEMENTS

This study was entirely funded by Médecins Sans Frontières-France.

