## Supplementary File for "COVID-19 in French Nursing Homes during the Second Pandemic Wave: A Mixed-Methods Cross-Sectional Study"

**Supplementary materials**

Appendix 1. Definitions

Appendix 2. Description of Study Participants and Interviews

Appendix 3. Interview Topic Guide for Caregivers and Residents

Appendix 4. Additional Descriptive Results

Appendix 5. Additional Kaplan Meier Curves

Appendix 6. Additional Cox Models

Appendix 7. Mixed Methods

### Appendix 1. Definitions

#### Autonomy Evaluation Score (Groupement Iso-Resources or GIR)

The GIR score is a measurement of autonomy loss based on a series of questions and observations, a team assesses a person’s level of dependency. In the Nursing Homes context, this evaluation is done by the coordinating physician upon admission of a new resident. The GIR score ranges from 1 to 6, from highest dependency (lowest level of autonomy) to lowest dependency.

GIR 1: includes elderly people confined to a bed or armchair, whose mental functions are seriously impaired and needing the continuous presence of caregivers.

GIR 2 reflects 2 categories:

-People confined to bed or a chair, whose mental functions are NOT totally impaired, and who need care for most activities of daily living;

-People whose mental functions are severely impaired but who have retained their ability to move around.

GIR 3 includes people who have retained their mental autonomy but who need help every day and several times a day to carry out everyday activities (getting up, going to bed, getting dressed, going to the bathroom, etc.).

GIR 4 reflects 2 categories:

-People in need of help to get up and go to bed, but able to move around the home on their own. They sometimes need assistance to dress and wash themselves;

-People who do not have motor impairment but need help with physical activities and meals.

Gir 5 groups together people who need occasional help with washing, preparing meals and cleaning.

Gir 6 refers to people who have fully retained their autonomy in the acts of daily life.

Reference for this definition (in French): <https://www.service-public.fr/particuliers/vosdroits/F1229#:~:text=La%20grille%20Aggir%20est%20utilis%C3%A9e,et%20sociales%2C%20dites%20activit%C3%A9s%20illustratives>

#### Average Weighted Autonomy Score (GIR Moyen Pondéré or GMP)

This score is calculated at the Nursing Home level and summarizes the overall level of residents’ dependency. Each resident requires X minutes of caregivers attention per day, X varying with the Autonomy score level (for ex. X=210 min for GIR 1; 88 min for GIR 4). The AWAS is then the average X residents need for the overall facility.

The higher the AWAS score, the more dependent the residents are. In other terms, the score is a proxy of the financial and human resources a Nursing Home can need and get: the higher the AWAS, the more resources the NH needs (higher staff-to-residents ratio, better equipment etc.).

Reference for this definition (in French): <https://assurance-dependance.ooreka.fr/astuce/voir/655507/gir-moyen-pondere>

#### Failure to thrive Syndrome^[1]^:

Specific to old age, this syndrome is defined by the rapid deterioration of the general state with anorexia, disorientation, social withdrawal, alongside a more or less directly expressed will to die, a passive give-up on life, an active refusal of care, of food. It evolves towards death in a few days to a few weeks. It is triggered by physical events (acute illnesses, surgery, trauma) or psychological events (death of a loved one, social isolation, hospitalization).

#### FFP2 (or N95 or KC95) Facemasks

The EN 149 standard defines performance requirements for three classes of [particle-filtering](https://en.wikipedia.org/wiki/Air_filter) half masks: FFP1, FFP2 and FFP3.

A FFP2 facemask filters at least 94% of airborne particles and has an internal leak rate of maximum 8%.

### Appendix 2. Description of Study Participants and Interviews

| **Participant characteristic** | | | | **Interview characteristic** | | |
| --- | --- | --- | --- | --- | --- | --- |
| Study n° | Function | | Sex | Duration | Type | Place |
| 1 | Directors | | Woman | 95 | individual | direction desk |
| 24 |  |  | Man | 119 | individual | direction desk |
| 31 |  |  | Man | 133 | individual | direction desk |
| 49 |  |  | Woman | 65 | individual | coordinator's desk |
| 10 | Coordinating doctors | | Woman | 45 | individual | coordinator's desk |
| 12 |  |  | Woman | 171 | individual | research desk |
| 48 |  |  | Woman | 55 | individual | infirmary |
| 2 | Coordinating nurses | | Woman | 71 | individual | direction desk |
| 13 |  |  | Woman | 32 | individual | coordinator's desk |
| 30 |  |  | Woman | 107 | individual | coordinator's desk |
| 56 |  |  | Woman | 68 | individual | coordinator's desk |
| 4 | Psychologists | | Woman | 35 | individual | coordinator's desk |
| 20 |  |  | Woman | 54 | individual | animators desk |
| 9 | Caregivers (internal permanent staff) | Assistant Nurse | Woman | 29 | individual | animators desk |
| 11 |  | Assistant Nurse | Woman | 28 | individual | collective room |
| 15 |  | Assistant Nurse | Woman | 37 | grouped (4 people) | animators desk |
| 16 |  | Assistant Nurse | Woman |  |  | animators desk |
| 17 |  | Animator | Woman |  |  | animators desk |
| 18 |  | Assistant Nurse | Woman |  |  | animators desk |
| 22 |  | Assistant Nurse | Woman | 21 | individual | infirmary |
| 23 |  | Nurse | Woman | 36 | individual | collective room |
| 27 |  | Assistant Nurse | Woman | 61 | grouped (2 people) | research desk |
| 28 |  | Assistant Nurse | Woman |  |  | research desk |
| 29 |  | Animator | Man | 67 | individual | research desk |
| 34 |  | Nurse | Woman | 46 | individual | research desk |
| 35 |  | Assistant Nurse | Woman | 48 | individual | collective room |
| 45 |  | Assistant Nurse | Woman | 55 | individual | infirmary |
| 46 |  | Assistant Nurse | Woman | 26 | individual | collective room |
| 51 |  | Nurse | Man | 49 | grouped (2 people) | infirmary |
| 5 | Caregivers (external staff) | Nurse | Woman |  |  | infirmary |
| 21 |  | Assistant Nurse | Woman | 25 | individual | rest room |
| 33 |  | Physiotherapist | Man | 37 | individual | private house |
| 44 |  | Physiotherapist | Man | 20 | individual | collective room |
| 47 | Other Staff | HRD manager | Woman | 56 | individual | coordinator's desk |
| 7 |  | Agent for Maintenance | Man | 48 | grouped (2 people) | maintenance desk |
| 8 |  | Agent for Maintenance | Man |  |  | maintenance desk |
| 25 |  | Cook | Woman | 38 | grouped (2 people) | kitchen |
| 26 |  | Cook | Woman |  |  | kitchen |
| 32 |  | Cleaner | Woman | 17 | individual | collective room |
| 52 |  | Cook | Woman | 12 | individual | kitchen |
| 3 | Residents |  | Woman | 63 | individual | bedroom |
| 6 |  |  | Woman | 28 | individual | collective room |
| 14 |  |  | Woman | 24 | individual | bedroom |
| 19 |  |  | Woman | 34 | individual | collective room |
| 36 |  |  | Woman | 95 | grouped (2 people) | collective room |
| 37 |  |  | Woman |  |  | collective room |
| 57 |  |  | Woman | 41 | individuel | bedroom |

### Appendix 3. Interview Topic Guide for Caregivers and Residents

| ***Questions to caregivers*** | ***Objectives*** |
| --- | --- |

| **1/** **Outbreak Chronology (Subjective Narratives)** | |
| --- | --- |
| -*Introduction*  -*Can you tell me how the epidemic has started and evolved in your institution?* | -identification of subjective phases  -qualification of temporalities  -information level assessment |
| -*What have been the most difficult times ?* | -assessing the impact of the epidemic |
| **2/Adaptations in Relation to the Crisis Management** | |
| *-The organisation of the NH was disrupted for a few weeks, how were practices reorganised in relation to : colleagues/ residents/ families ?* | -description of crisis effect |
| *-Have you received any external aid? In what areas?* | -networks, actors’ schemes |
| -*What permitted a return to normal activity?*  -*What could be enhanced in terms of crisis management?* | -return to normal activity |
| **3/** **Individual Experience of the Second Pandemic Wave** | |
| -*How did you become [function: a director, coordinating physician, nurse, assistant nurse, etc.] ?* | -socio-demographic profile  -University and professional trajectory |
| -*Did you receive any help in your work position?* | -networks, actors’ schemes  -collective participation  -description of isolation, understaffing |
| -*As a [function], how did you experience this period?* | -ethical questionings  -individual variables (personal, family, emotional) |

| ***Questions to residents*** | ***Objectives*** |
| --- | --- |

| **1/Outbreak Chronology (Subjective Narratives)** | |
| --- | --- |
| -*Introduction*  -*Can you tell me about the period of COVID in the NH?*  *-What were the differences compared to other periods in the past year?* | -identification of subjective phases  -qualification of temporalities  -information level assessment |
| -*What have been the most difficult times ?* | -assessing the impact of the epidemic |
| **2/Adaptations in Relation to the Crisis Experience** | |
| *-Have you been contaminated with COVID? Have you been hospitalized?*  *-Have you been particularly worried about this disease? (isolation, containment)* | -situation and positioning of the individual in relation to the epidemic  -description of crisis effect |
| *-Did you see other neighbours/friends of the NH?*  *-Did you see relative/ family members outside the NH? In the NH?*  *-Were there any activities?* | -networks, actors’ schemes  -links with the outside world |
| *-Were you moved during COVID?*  *-What do you think about the organisation of the NH staff during COVID? What could have been improved?* | -identification of novelty  -return to normal activity |
| **3/Individual Experience of the Second Pandemic Wave** | |
| -*In what year were you born? In what year did you enter the NH?*  *-Before the NH, what did you do? Where did you live?* | -geographic trajectory before the NH  -trajectory within NH  -socio-demographic profile  -University and professional trajectory |
| *-(in normal times) Do you prefer to stay in your room? To participate in group activities?*  *-Have you received any support apart from the assistant nurses/ nurses? In what areas?*  *-Did your attending physician come?*  *-Have you had contact with your relatives?* | -networks, actors’ schemes  -collective participation  -description of isolation |
| *-Do you have family members in the area, elsewhere?*  *-Do you have relatives who have had COVID?* | -individual variables (personal, family, emotional) |

### Appendix 4. Additional Descriptive Results

Table1. General and epidemiological characteristics of 22 nursing homes (aggregated data)

| Facility Data | N | Mean | Std Dev | Min | Max |
| --- | --- | --- | --- | --- | --- |
| Number of beds | 22 | 80.32 | 19.1 | 50 | 121 |
| Average Weighted Autonomy Score | 20 | 775 | 44.7 | 686 | 870 |
| Time to FFP2 use (days) | 22 | 8.7 | 8.7 | 0 | 28 |
| Time to MSF Intervention (days) | 22 | 18.9 | 9.7 | 5 | 37 |
| Staff-to-residents Ratio | 22 | 0.81 | .14 | .53 | 1.09 |
| Number of Staff | 22 | 61.3 | 18.9 | 32 | 109 |
| Number of Residents | 22 | 74.7 | 17.0 | 44 | 106 |
| COVID-19 episode duration (days) | 22 | 37.8 | 14.9 | 6 | 81 |
| Attack Rate in Staff (%) | 22 | 38.1 | 18.4 | 23.8 | 71.4 |
| Attack Rate in Residents (%) | 22 | 65.6 | 20.0 | 13.8 | 96.0 |
| Case Fatality Rate in residents (%) | 22 | 19.4 | 10.0 | 0 | 39.7 |

Comorbidities vs FTTS (Fischer Exact Test p-value= 0.051)

|  | Failure to thrive syndrome | |  |
| --- | --- | --- | --- |
| Comorbidities | No  410 (77.2%) | Yes  121 (22.8%) | Total  531 |
|  | N (row %) | N (row %) |  |
| None | 159 (71.9%) | 62 (28.1%) | 221 |
| 1 | 116 (85.3%) | 20 (14.7%) | 136 |
| 2 | 84 (79.2%) | 22 (20.8%) | 106 |
| 3 | 37 (75.5%) | 12 (24.5%) | 49 |
| >=4 | 14 (76.7%) | 5 (26.3%) | 19 |

Pearson pairwise correlation matrix for Average Weighted Autonomy Score, Nursing Home Size and Staff-to-Resident Ratio (continuous) :

|  | AWAS | Number of residents |
| --- | --- | --- |
| AWAS | 1.0000 |  |
| Number of residents | 0.5356* | 1.0000 |
| Staff-to-Resident Ratio | 0.6617* | 0.1776* |

*p-value < 0.05

AWAS vs Staff -to-Resident Ratio (categories): Fischer Exact Test p-value< 0.001)

|  | Staff to Resident Ratio (Cat.) | | |  |
| --- | --- | --- | --- | --- |
| AWAS (cat) | Low  (<0.8) | Medium  (0.8-0.9) | High  (>=0.9) | *Total* |
| Low (<=750) | 154 | 26 | 46 | *226* |
| Medium (750-800) | 0 | 84 | 24 | *108* |
| High (>=800) | 0 | 80 | 171 | *251* |
| *Total* | *154* | *190* | *241* | *585* |

AWAS vs Nursing Home Size (categories): Fischer Exact Test p-value< 0.001)

|  | Nursing Home Size (cat) | | |  |
| --- | --- | --- | --- | --- |
| AWAS (cat) | <70 res. | 70-90 | >=90 | *Total* |
| Low (<=750) | 170 | 56 | 0 | *226* |
| Medium (750-800) | 108 | 0 | 0 | *108* |
| High (>=800) | 27 | 115 | 109 | *251* |
| *Total* | *305* | *171* | *109* | *585* |

Staff to Resident Ratio vs Nursing Home Size (categories): Fischer Exact Test p-value < 0.001)

|  | Nursing Home Size (cat.) | |  |
| --- | --- | --- | --- |
| Staff to Resident Ratio (Cat.) | <70 res. | >=70 res. | *Total* |
| Low (<0.8) | 98 | 56 | *154* |
| Medium (0.8-0.9) | 101 | 89 | *190* |
| High (>=0.9) | 70 | 171 | *241* |
| *Total* | *269* | *316* | *585* |

Pearson correlation matrix for Time to FFP2 use, Time to MSF intervention and duration of COVID-19 episode

|  | Time to FFP2 use | Time to MSF intervention |
| --- | --- | --- |
| Time to FFP2 use (cont.) | - |  |
| Time to MSF intervention (cont.) | 0.0989 | - |
| Duration of COVID-19 episode (cont.) | 0.5523* | 0.5250* |

*p-value < 0.05

Time to FFP2 use vs Time to MSF (categories): Fischer Exact Test p-value < 0.001)

|  | Time to MSF intervention (cat) | | |  |
| --- | --- | --- | --- | --- |
| Time to FFP2 use (cat) | Short (<10 days)  78 (13.3%) | Medium (10-20 days)  326 (55.7%) | Long (>20 days)  181 (30.9%) | *Total* |
| Instant.(<=1day) | 8 (6.8%) | 54 (45.8%) | 56 (47.4%) | *118* |
| Late (2-7 days) | 70 (41.4%) | 53 (31.3%) | 46 (27.2%) | *169* |
| Very Late (>=7 days). | 0 | 219 (73.5%) | 79 (26.5%) | *298* |

### Appendix 5. Additional Kaplan-Meier Curves – full list (for Log Rank Tests results, see Table 1 in main manuscript)

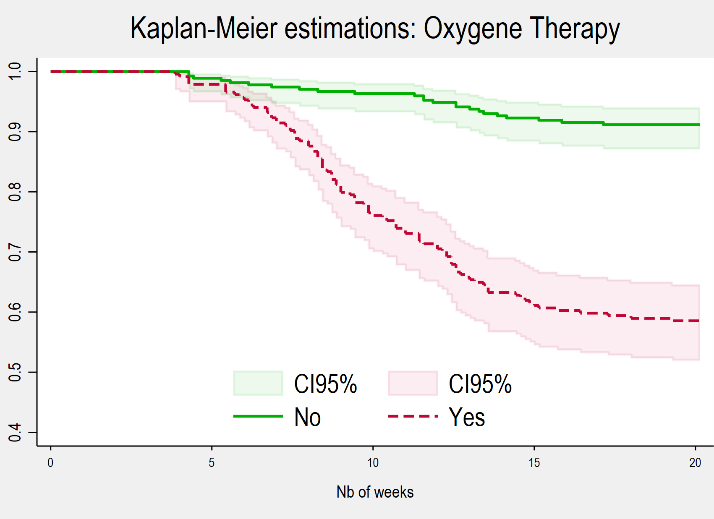

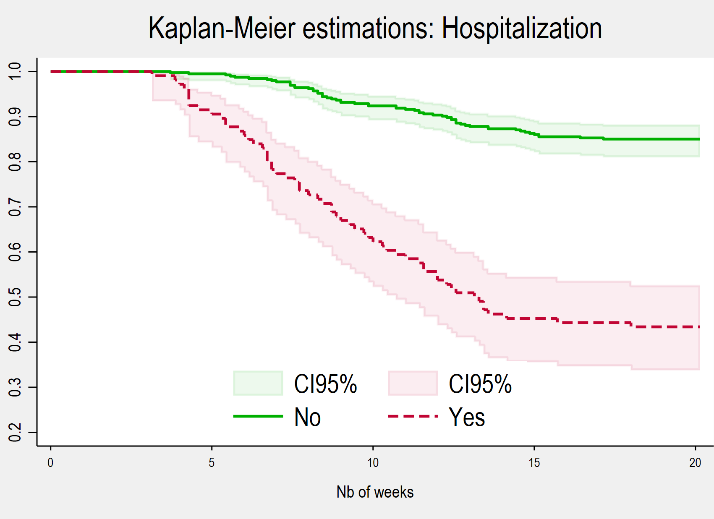
Individual Data (Linelist)

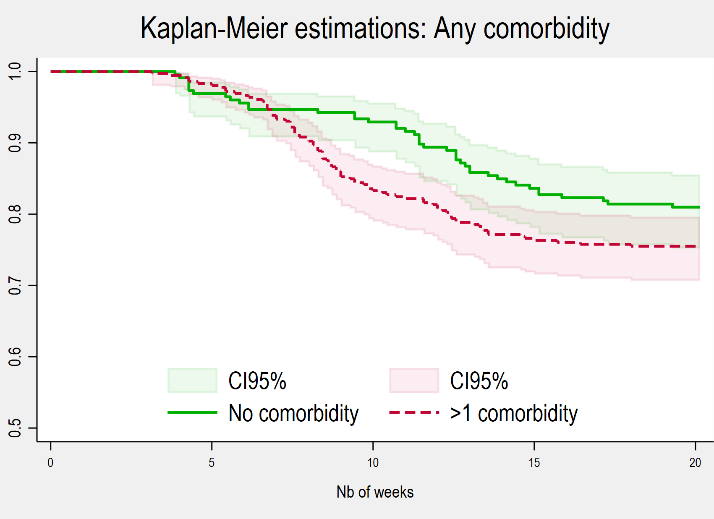

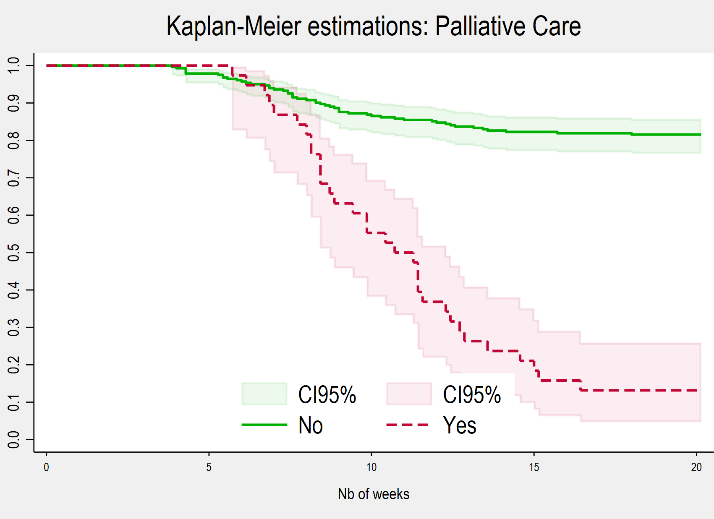

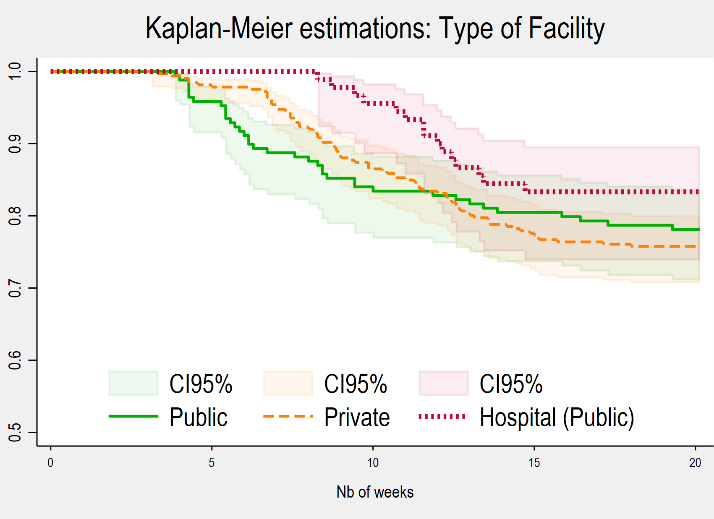
Facility Data (aggregated)

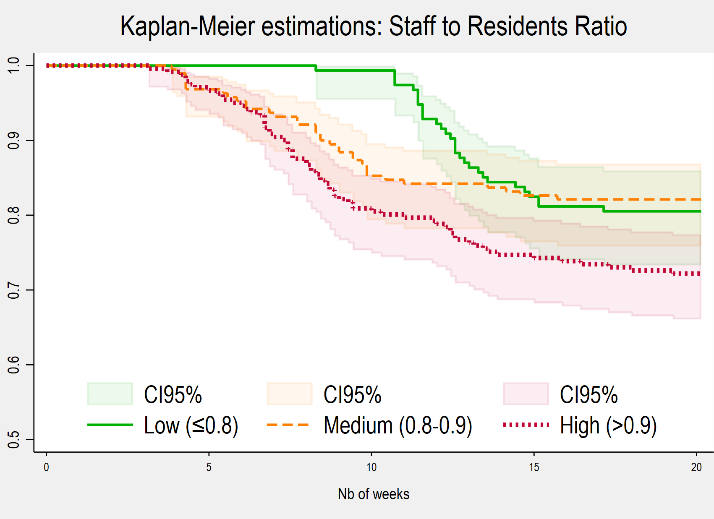

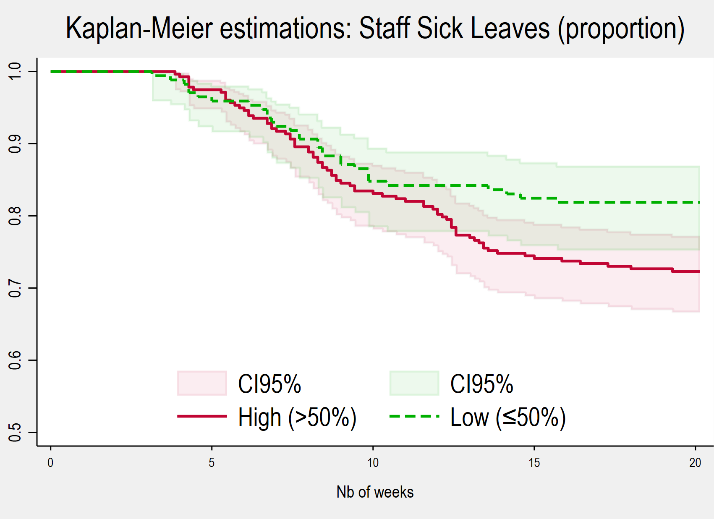

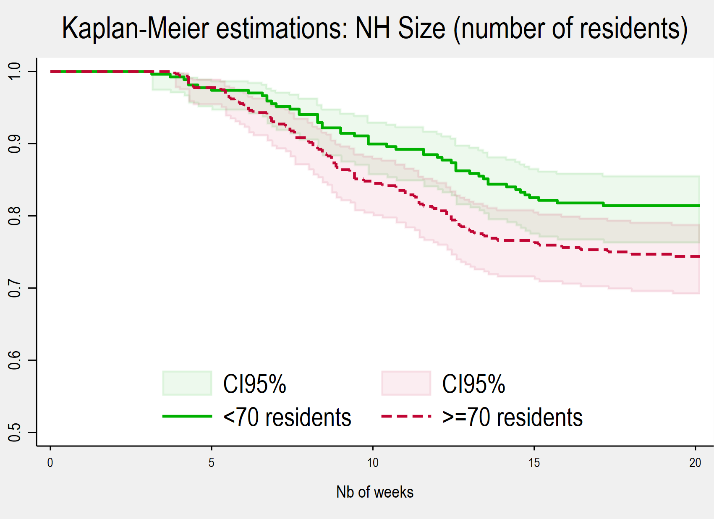

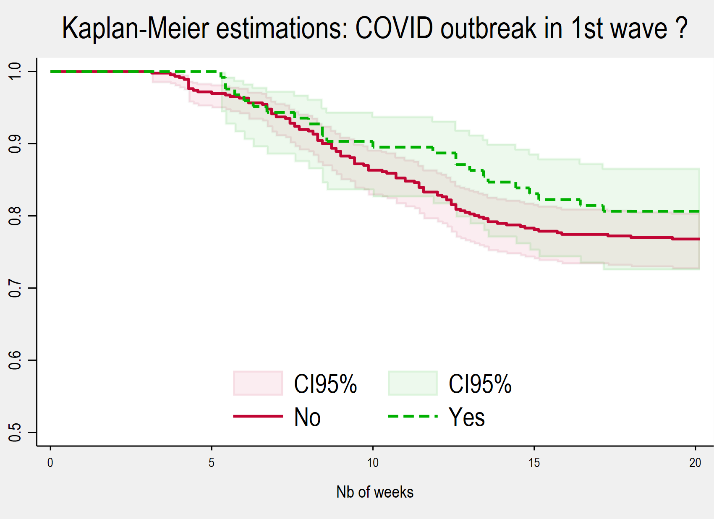

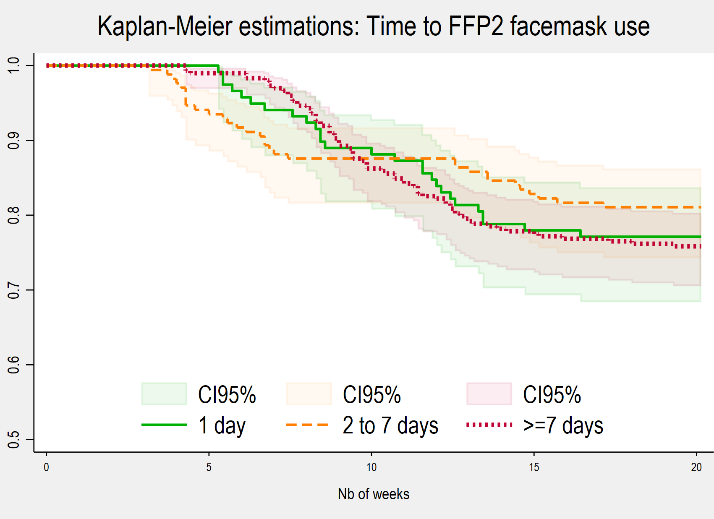

### Appendix 6. Additional Cox models (Sensitivity Analysis)

Model 1. Only individual data with ‘obvious’ covariates (hospitalization, palliative care etc)

| **VARIABLES** |  | **Adjusted Hazard Ratio** | **CI95** | **p-value** |
| --- | --- | --- | --- | --- |
| Age | Continuous | 1.00 | 0.97 - 1.03 | 0.921 |
| Autonomy Score | 2 vs 0 | 0.89 | 0.48 - 1.66 | 0.715 |
|  | 3 vs 0 | 0.53* | 0.26 - 1.09 | 0.085 |
|  | >=4 vs 0 | 0.40* | 0.14 - 1.11 | 0.078 |
| Gender | M vs F | 1.62* | 0.93 - 2.84 | 0.088 |
| Comorbidities | 1 vs 0 | 1.83 | 0.46 - 7.29 | 0.391 |
|  | 2 vs 0 | 1.64 | 0.42 - 6.39 | 0.473 |
|  | 3 vs 0 | 2.02 | 0.48 - 8.53 | 0.340 |
|  | >=4 vs 0 | 2.73 | 0.50 - 15.06 | 0.248 |
| Hospitalization | Y v N | 4.19*** | 2.53 - 6.91 | 0.000 |
| Oxygene Therapy | Y v N | 3.08*** | 1.42 - 6.64 | 0.004 |
| Palliative Care | Y v N | 3.09*** | 1.69 - 5.63 | 0.000 |
| Failure-to-thrive Syndrome | Y v N | 3.22** | 1.14 - 9.09 | 0.027 |
| Interaction terms | Comorb=1#FTTS=1 | 0.84 | 0.17 - 4.05 | 0.824 |
|  | Comorb=2#FTTS=1 | 0.72 | 0.18 - 2.90 | 0.648 |
|  | Comorb=3#FTTS=1 | 0.77 | 0.14 - 4.22 | 0.763 |
|  | Comorb=4#FTTS=1 | 1.02 | 0.13 - 8.13 | 0.982 |
|  | Hospitalization=1#Oxygene=1 | 0.39 | 0.04 - 3.31 | 0.389 |
|  | Oxygene=1#Palliative=1 | 0.14# | 0.07 - 0.28 | 0.000 |

\# interaction term significant > oxygene effect amplified by palliative care effect

| AIC | BIC |
| --- | --- |
| 696.215 | 722.30 |

Information Criteria (model selection)

Model 2. Only individual data with detailed comorbidities

| **VARIABLES** |  | **Adjusted Hazard Ratio** | **CI95** | **p-value** |
| --- | --- | --- | --- | --- |
| Age | Continuous | 1.00 | 0.97 - 1.03 | 0.921 |
| Autonomy Score | 2 vs 0 | 0.89 | 0.48 - 1.66 | 0.715 |
|  | 3 vs 0 | 0.53* | 0.26 - 1.09 | 0.085 |
|  | >=4 vs 0 | 0.40* | 0.14 - 1.11 | 0.078 |
| Gender | M vs F | 1.79* | 1.16 - 2.74 | 0.008 |
| Diabetes | Y v N | 2.81** | 1.17 - 6.76 | 0.021 |
| Denutrition | Y v N | 2.54 | 0.55 - 11.82 | 0.235 |
| Dementia | Y v N | 0.91 | 0.40 - 2.08 | 0.822 |
| Cardiovascular Disease | Y v N | 1.24 | 0.75 - 2.06 | 0.409 |
| Cancer | Y v N | 0.96 | 0.42 - 2.19 | 0.919 |
| Obesity | Y v N | 1.37 | 0.46 - 4.04 | 0.571 |
| Respiratory Disease | Y v N | 0.68 | 0.22 - 2.15 | 0.514 |
| High Blood Pressure | Y v N | 0.91 | 0.56 - 1.48 | 0.712 |
| Failure-to-thrive Syndrome | Y v N | 4.79*** | 1.52 - 15.06 | 0.007 |
| Interaction terms | AES=2#FTTS=1 | 2.54 | 0.80 - 8.10 | 0.114 |
|  | AES=3 # FTTS=1 | 3.21 | 0.78 - 13.16 | 0.105 |
|  | AES=4# FTTS=1 | 4.94 | 0.51 - 48.01 | 0.169 |
|  | FTTS=1#Diabetes=1 | 0.20# | 0.04 - 1.05 | 0.057 |
|  | FTTS=1#Denutrition=1 | 0.15# | 0.03 - 0.86 | 0.033 |
|  | FTTS=1#Dementia=1 | 1.21 | 0.44 - 3.31 | 0.717 |
|  | Diabetes=1# Denutrition=1 | 0.40 | 0.03 - 4.61 | 0.461 |
|  | Diabetes=1# Dementia=1 | 0.75 | 0.16 - 3.38 | 0.703 |
|  | Denutrition =1# Dementia=1 | 1.24 | 0.24 - 6.34 | 0.792 |
|  | HBP=1#Cardiovasc=1 | 1.19 | 0.45 - 3.18 | 0.723 |

\## interaction term significant > FTTS effect amplified by Denutrition effect and by diabetes effect

Information Criteria (model selection)

| AIC | BIC |
| --- | --- |
| 770.399 | 803.8226 |

Model 3. Individual and structural data with Staff-to-Resident Ratio and NH Size instead of AWAS

| **VARIABLES** |  | **Adjusted Hazard Ratio** | **CI95** | **p-value** |
| --- | --- | --- | --- | --- |
| Age | Continuous | 1.00 | 0.99 - 1.01 | 0.635 |
| Autonomy Score | 2 vs 0 | 0.70 | 0.31 - 1.58 | 0.388 |
|  | 3 vs 0 | 0.40** | 0.17 - 0.95 | 0.038 |
|  | >=4 vs 0 | 0.23*** | 0.08 - 0.66 | 0.006 |
| Gender | M vs F | 1.78** | 1.12 - 2.81 | 0.014 |
| Comorbidities | 1 vs 0 | 1.28 | 0.52 - 3.16 | 0.590 |
|  | 2 vs 0 | 1.20 | 0.63 - 2.25 | 0.580 |
|  | 3 vs 0 | 1.40 | 0.51 - 3.82 | 0.517 |
|  | >=4 vs 0 | 1.67 | 0.51 - 5.46 | 0.396 |
| Failure-to-thrive Syndrome | Y v N | 4.07*** | 1.94 - 8.54 | 0.000 |
| Presence of a physician | Half Time vs None/Absent | 0.26*** | 0.13 - 0.53 | 0.000 |
|  | Full Time vs None/Absent | 0.26*** | 0.10 - 0.64 | 0.004 |
| Time to FFP2 use (in days) | continuous | 1.01 | 0.95 - 1.07 | 0.681 |
| Staff to Resident Ratio | continuous | 1.17 | 0.84 - 1.35 | 0.586 |
| NH Size (number of residents) | continuous | 1.03 | 0.93 - 1.14 | 0.545 |
| Staff Attack Rate (%) | continuous | 2.18 | 0.29 - 16.49 | 0.450 |
| Interaction terms | AES=2#FTTS=1 | 2.30# | 0.91 - 5.78 | 0.077 |
|  | AES=3#FTTS=1 | 2.93# | 0.95 - 9.05 | 0.061 |
|  | AES=4#FTTS=1 | 4.80# | 1.16 - 19.92 | 0.031 |
|  | NR_Ratio#NH Size | 0.95 | 0.83 - 1.08 | 0.402 |

\## interaction term significant > FTTS effect amplified at each level of AES effect

| AIC | BIC |
| --- | --- |
| 1172.544 | 1227.964 |

Information Criteria (model selection)

**Appendix 7. Mixed Methods**

**Multidisciplinary Research and Collective Protocols**

Both quantitative and qualitative data collection stem from an iterative reflexive process within the interdisciplinary research team, comprising: a social geographer (M.D.) and a public health expert (S.F.) present on the fieldwork (both are PhD female researchers employed at Epicentre for this research project and trained in fieldwork methods with vulnerable populations in crisis contexts); a lead epidemiologist (T.R.), a medical doctor (T.L.), a MSF project coordinator (C.M.), a nurse (C.S.) and a psychologist (M.T.) partly present on the research fieldwork; and two coordinating epidemiologists working at Epicentre (E.G. and K.P.).

During the exploratory phase (from 1st December 2020 to 22 January 2021), several focus groups were organised within the MSF-team, in order to define the research objectives, the strategy for selecting research sites for qualitative analysis, and key resource interlocutors. Regular informal and semi-structured meetings with MSF nurses, and analytical reading of their monitoring reports from emergency interventions, both helped in drafting the research protocol and fieldwork priorities. The interview topic guide (Appendix 3) and a checklist for systematic observation were conceived by M.D. and commented by MSF coordinators on the fieldwork (C.M., T.L., C.S.). Throughout this collective process and preliminary analyses, the public health expert (S.F.) conceived a database. The social geographer (M.D.) and the public health expert (S.F.) both visited a few nursing homes with the MSF coordinators before formally beginning the research.

On the fieldwork (from 22 January to 26 February 2021), the public health expert (S.F.) collected most epidemiological data, as well as individual data for retrospective linelist analyses. The social geographer (M.D.) gathered most qualitative data, including direct observation notes and semi-structured interviews, for 4 nursing homes. However, the two fieldwork researchers worked together narrowly. They managed together first contacts with the directors and/or coordinating physicians of the studied nursing homes, they visited together 2 nursing homes out of the 4 comprised in the qualitative study, they compared their results on a daily basis and organised their data commonly.

In the phase of reporting (from the 1^st^ March to the 21^st^ April 2021), an internal report was written and sent for proofreading to the interdisciplinary research team. In the following month, a synthetic report was written. Corrections after proofreading were incorporated in May and June 2021. The final reports were sent to interviewees in June and September 2021 for comments. Only few feedbacks were received, mostly on formal aspects.

**Statistical Methods**

We first performed a descriptive analysis of the data collected by the MSF team from NH managers: facility-level information and linelists (COVID-19 cases among residents). We crossed several factors with the resident’s final status and computed Kaplan-Meier estimations of the probability of dying from COVID-19 in parallel with univariate Cox model for each factor. Log-Rank Test was used to assess potential association of each factor with death. Date of entry in the study was set to October 25th, 2020 (date of the new prevention measured announced by the French government and start of the second wave in France). Date of exit was set to March 15^th^, 2021 (official end of the study), in case of death, to the exact date of death (if available).

We then explored the probability of dying from COVID-19 according to the factors identified in the univariate analysis with Cox models (multivariate analysis).

The challenge with multivariate analyses stems from the fact that various individual and structural factors may possibly be associated, and some of them can also be considered as confusion factors.

Variables reflecting a notion of temporality, such as the time to FFP2 use and time to MSF intervention or attack rate among residents/staff and duration of the COVID-19 episode may be correlated and may not all be included in a single model. In a similar fashion, proportion of sick leaves in staff and characterization of the physician presence are obviously correlated.

We thus built several Cox models depending on the factors we wanted to include. We decided to control for age, autonomy level and gender in all models.

One model analyzed detailed comorbidities (cancer, high blood pressure etc.) in order to highlight potential risk/protective factors of death. Another model analyzed a summary of comorbidities (absence/presence of >1 comorbidity or total number of comorbidities).

We then tested alternate models analyzing either quantitative factors as continuous variables or transformed versions of the same factors as categorical variables (using cutoffs).

Choice of variables to finally retain in each model followed a classical Stepwise selection process, starting from a model gathering factors for which p-values (association with mortality according to Log-Rank Test) were < 0.3.

We have taken into account the many interactions that come into play between several factors: hospitalization with oxygene therapy and/or palliative care, interrelated comorbidities (high blood pressure with cardiovascular disease, obesity and diabetes etc.), Failure-to-thrive syndrome with comorbidities, AES with comorbidities or Failure-to-thrive syndrome, time-reflecting factors (as seen previously: time to FFP2 use, time to MSF intervention, duration of COVID episode, attack rates).

We fitted mixed-effects three-level random-slope exponential survival models. To account for individual heterogeneity, we included a random effect at the individual level, and to account for clustering, we included a random effect at the nursing home level (individuals are nested within each nursing home). Robust Standard Errors were computed and presented (clustered at the highest level in the multilevel model, here the nursing home).

**Qualitative Study Context**

Qualitative methods are interrelated with the context of the research. The interviews followed MSF interventions and epidemic peaks in the NH. The relative respite after the outbreaks favoured data collection: interviewees were more eager to give time to the study than during the outbreaks’ peaks.

The major interests expressed in the research topics were that the participants were thankful to MSF teams, saw research as a way to step back from the traumatic experience of high fatality cases in their NH, to express a silenced point of view, or to contribute to general knowledge on the issue of COVID-19.

The access to the fieldwork through MSF helped organising rapidly a confident environment for the interviews to take place, since MSF support was mostly very welcomed and appreciated, as participants reported to the lead investigator (M.D.). For the same reason, the lead investigator could be considered as a member of MSF, which could have resulted in possible biases; therefore, the distinction between MSF interventions and Epicentre research had to be underlined before each interview.

Objectives, risks and benefices of the study were explained thanks to information letters for participation in the study and informed consent forms that were read and signed before the interviews. Each participant was informed that participation to the study is free, can be interrupted without justification and at any time without consequences. Each participant had a time for thinking, questioning and possibly obtaining explanations from the interviewer.

Methods of anonymization et confidentiality were applied for all participants, following the good practices identified by the Institute for Human and Social Research of the French National Center for Scientific Research (InSHS-CNRS).

1. Palmer RM. 'Failure to thrive' in the elderly: diagnosis and management. Geriatrics. 1990 Sep;45(9):47-50, 53-5. PMID: 2204587.
